# Maternal and child healthcare-seeking among victims of violence in armed conflict: A quasi-experimental study in northeast Nigeria∗

**DOI:** 10.1101/2025.03.11.25323760

**Authors:** Max Schaub, Gbadebo Collins Adeyanju, Aisha Aliyu Abulfathi, Musa Muhammad Bello, Lennart Kasserra, Aminatu Ayaba Kwaku, Muhammad Ibrahim Jalo, Ahmad Mahmud, Pia Schrage, Rabiu Ibrahim Jalo, Liliana Abreu

## Abstract

**Introduction:** Armed conflict severely impacts health, with indirect deaths often exceeding direct casualties 2–4 times, disproportionately affecting women and children. Although the magnitude of these effects is well-documented, the mechanisms driving them remain insufficiently understood. This study shifts the focus from supply-side factors, such as the destruction of infrastructure, to demand-side processes, particularly healthcare-seeking behavior, and from broader conflict exposure to individual-level violent victimization.

**Methods:** Data come from a representative survey (n = 3,006) of caregivers of young children in northeastern Nigeria, a region heavily affected by armed insurgency. Unlike previous studies, our survey included dedicated measures of victimization, health-seeking outcomes, and potential mediating factors within a single instrument, enabling precise measurement and analysis. To compare maternal and child healthcare-seeking behavior between victimized and non-victimized caregivers, we employed a quasi-experimental observational design using propensity-score matching on demographic and contextual characteristics. Causal mediation analysis was then used to identify the mechanisms linking victimization to health behaviors.

**Results:** Victimization was widespread, with 21% of respondents (n=651) having experienced a severe form of violence in the past 3 years. While maternal healthcare-seeking behavior appeared unaffected, victimization significantly reduced healthcare-seeking for child health. Children of victimized caregivers were markedly less likely to be fully immunized (Odds ratio (OR): 0.43, p<0.001) and to receive care at government health facilities (OR: 0.23, p<0.001). This decline was primarily driven by increased distrust in the health system (PM: 0.42–0.70, p<0.05), stemming from negative experiences during the conflict, particularly health worker absenteeism and victimization by state security forces.

**Conclusion:** Addressing fear and mistrust is key to improving healthcare-seeking in conflict-affected populations. Efforts should focus on providing security for government-run health facilities, reducing violence against civilians by state security forces, and restoring trust in healthcare and state institutions. Future research should explore effective strategies for achieving these objectives.

**What is already known?:**

- Armed conflict has severe negative consequences for maternal and child health, yet the underlying mechanisms remain understudied.
- Recent research has called for a greater focus on how exposure to violence shapes the demand for health services and healthcare-seeking behavior—a gap this study addresses with a representative survey (n = 3,006) in northeastern Nigeria.

**What are the new findings?:**

- Rather than approximating exposure through proximity to conflict, this study directly measures individual-level victimization and examines potential mediating mechanisms with high precision.
- 21% of respondents reported severe victimization; these individuals showed markedly lower child immunization rates (OR = 0.43, p < 0.001) and were less likely to use government health facilities (OR = 0.23, p < 0.001).
- These differences are driven by mistrust of healthcare workers, predicated on absenteeism and prior victimization by state security forces.

**What do the new findings imply?:**

- Preventing further erosion of trust likely requires reliable health service delivery and restraint by security forces.
- To improve child health outcomes, trust-building in healthcare and state institutions should be integrated in recovery programs.

## Introduction

Studies of armed conflict and health have documented the devastating negative health effects of exposure to violence. Estimates suggest that indirect deaths from conflict (e.g., due to malnutrition and diseases) often exceed direct casualties by a factor of two to four [1–3], with women and children disproportionately affected [3–5]. While these broad patterns are well documented, gaps remain in our understanding of why violence has such detrimental health consequences. The best-documented explanations focus on supply-side factors: armed conflict is accompanied by the large-scale destruction of critical infrastructure, leaving affected populations without essential services and vulnerable to the spread of infectious diseases [6–9]. In addition, health workers frequently flee conflict-affected areas, further limiting the availability of medical services [7, 10–13]. Looking beyond such supply-side dynamics, recent research has shifted the focus to demand-side factors, particularly how exposure to violence affects healthcare-seeking behavior, defined as utilization of health services in armed conflict settings [12, 14–18]. Exposure to violence can induce fear, scholars have argued, leading individuals to reduce their movement and limit their pursuit of maternal and child healthcare [17–19]. Moreover, individuals living in conflict-affected regions often report lower trust in government institutions, a key predictor of healthcare utilization [17, 20–22].

In this regard, Nigeria represents a particularly relevant case. The country faces multiple forms of armed conflict, including the Boko Haram insurgency in the Northeast, recurrent clashes between farmers and herders in the Middle Belt, separatist agitation in the Southeast, and criminal banditry in the Northwest [23, 24]. These conflicts have had profound repercussions for the health system, ranging from the destruction of facilities and the displacement of health workers to interruptions in immunization programs and disease surveillance [25, 26]. In the Northeast, where our study is situated, healthcare capacity has been particularly strained, with many communities reliant on humanitarian organizations whose reach remains inconsistent due to insecurity [27, 28]. Existing research on Nigeria underscores the severity of these challenges: attacks on health services have contributed to elevated maternal and child mortality, malnutrition, and outbreaks of preventable diseases such as cholera and measles [29, 30]. Studies also document the psychological toll of conflict, including high levels of post-traumatic stress and depression among displaced populations [31, 32].

Against this backdrop, the present study contributes to this literature, but shifts the focus from broader exposure to violence—typically measured as proximity to attacks—to individual-level violent victimization. Even though victimization is one of the most direct consequences of exposure to armed conflict, its impact on healthcare-seeking has not been explicitly assessed. In conflict settings, large numbers of individuals suffer from attacks themselves or observe family members being killed or otherwise harmed. Our study systematically measures such instances of victimization and assesses their relationship with healthcare-seeking behavior. Additionally, we advance existing research by explicitly identifying and modeling potential pathways through which victimization translates into reduced healthcare-seeking, focusing on trauma, fear, and mistrust. It is well established that victimization can lead to persistent trauma, with survivors exhibiting significantly higher rates of post-traumatic stress symptoms, even decades after exposure [33–35]. As a consequence of trauma, victims may under-utilize healthcare services [36, 37]. It is also plausible that violent victimization may increase fear of further victimization, discouraging individuals from leaving their homes to seek healthcare for themselves, their partner, or their children [19, 37, 38]. Similarly, victimization can erode trust in healthcare institutions. This may be because of negative experiences with healthcare services during the conflict or a generalization of mistrust from one state institution (e.g., the security forces that failed them) to other state institutions, such as government-provided healthcare. Distrustful individuals will stop using health services, e.g., by refusing to vaccinate their children [39, 40], or they will seek non-government-run alternatives [41, 42]. In order to consistently track these potential mediation effects, our study included measures for all of these dimensions in the same survey instrument.

## Methods

### Data

Data come from a representative household survey (n=3,006) conducted in August and September 2024 in the three northeastern Nigerian states of Adamawa, Borno, and Yobe (see Figure S1 in the Supplementary Information (SI) for a graphical overview). The region has been severely affected by an armed insurgency for the last 15 years [48]. And while the security situation has stabilized somewhat in recent years, sporadic attacks on government and civilian targets persist, including raids by armed extremists on villages and attacks on hospitals [49]. In each state, two local government areas (LGAs, comparable to counties) were randomly selected, weighting by population size. Within each LGA, three political wards (out of 10 to 20) were chosen using the same weighting approach. Due to security constraints, three wards had to be replaced during data collection with random replacements. The primary sampling units (PSUs) were determined by a population-weighted draw of 500*×*500m grid cells, which generated interviewer starting points proportional to population size. Household recruitment followed a random walk with a 2:1 skip pattern. Sampling weights were generated to adjust for any over-or under-representation of PSUs in the final sample, ensuring unbiased summary statistics. Additional information about the survey, in line with the STROBE reporting guidelines [46], is provided in Tables S1–S3 in the SI, and replication data and code are available on OSF [47].

The target population for our survey were parents (men and women) of young children. In each household, a single person was chosen for interview. To be eligible for the interview, a person had to be the primary caregiver of a child 5 years of age or younger. Interviews were conducted by trained interviewers in the local language (Hausa) using a pre-tested questionnaire. Participants were informed in advance of the sensitive nature of some of the questions included in the survey, and were informed that they could end the survey or skip individual questions at any time. Only individuals who gave their explicit informed consent were interviewed. Participants provided demographic information, answered questions about maternal and child health, and reported on experiences of violence and attitudes toward government and healthcare institutions. For child health, we focused on the youngest child in the family, aged at least one year. For maternal health, we asked female respondents to report on their own experiences and male respondents to report on their partners’ experiences. The survey and data collection protocol were approved by the National Health Research Ethics Committee of Nigeria (NHREC; Approval No. NHREC/01/01/2007-14/01/2024), by the respective authorities of the three states, and by the University of Konstanz, Germany (IRB Statement 13/2024).

### Patient and public involvement

The research project built on knowledge gained from qualitative fieldwork with focus group participants and key informants (community leaders and healthcare workers). These interviews provided important input for defining the final set of questions included in the survey. The data and findings will be disseminated through a series of workshops, the first of which was held in Kano, Nigeria, in January 2025, and which are open to specialists from Nigerian ministries, nongovernmental organizations (NGOs), and journalists.

### Study design and statistical approach

For the analysis, we adopted a quasi-experimental propensity score matching approach. We matched individuals who reported being victimized in the past three years with non-victimized individuals using a 1:1 matching algorithm. For statistical modeling, we considered both conditional and unconditional logistic regression. Conditional logit relies only on within-pair variation, offering a rigorous approach to causal inference, but it discards a substantial portion of the data and limits flexibility for mediation analysis. The causal mediation framework we employ [63] requires an outcome model that can accommodate continuous and multiple mediators. Because conditional logit removes stratum-specific intercepts, it complicates estimation of mediated effects and precludes the inclusion of mediators that do not vary within pairs. By contrast, unconditional logistic regression allows us to estimate the outcome model required for mediation analysis [64]. We therefore opted for unconditional models as our default specification, applied to matched data with adjustment for the matching variables and clustering of standard errors at the matched-pair level [65, 66]. Robustness checks using conditional logistic regression are reported in Section H of the SI; substantively, both approaches yield very similar estimates.

We matched on the demographic characteristics of the respondent (sex, age, education, number of children) and of the child in question (sex, age) to shield against potential confounding along these dimensions. For example, age and sex may be related to both the likelihood of victimization and healthcare-seeking patterns. Second, to isolate the *demand* for healthcare, we also accounted for determinants of healthcare supply. Specifically, we included the self-reported walking distance (in minutes) to the nearest clinic and the remotely measured geographic distance between the interview locations and georeferenced clinic and hospital locations [52]. We further included several health indicators in our matching function to capture the relative quality of services, namely diphtheria-tetanus-pertussis (DPT3) and measles vaccination coverage, malaria and malnutrition prevalence, and the under-five mortality rate [53–57]. A full description of the included contextual covariates can be found in Table S14 in the SI. These indicators were recorded at the 500*×*500m grid cell level, the same scale used for sampling. Finally, we included covariates that are known determinants of armed violence, notably population density, accessibility (measured as the distance to the nearest major road), and state reach (measured as the distance to the regional capital) [58]. All time-varying context-level covariates were measured pre-treatment, between 2015 and 2019, i.e., before the start of the recording-period for victimization. Matching was performed on the propensity score, estimated by means of a probit model regressing the victimization indicator on the full set of controls, and a caliper set to 0.2 [59]. This procedure matched the full set of victimized individuals (n=651) to the same number of non-victimized control-group individuals, for an effective sample of n=1,302 used in most of the analyses.

Our models relate violent victimization to indicators of healthcare-seeking for maternal and child health. Violent victimization was assessed using a survey question asking whether the respondent or a household member had been “directly affected by organized violence” in the past three years. Organized violence was defined as violence perpetrated by “members of an organized group, such as an extremist group or armed bandits.” Respondents were further instructed that being “affected” meant being “either the target or witness of violence.” Following this question, we asked whether respondents would mind sharing what had happened to them or their household member. We coded individuals as violently victimized if they reported that they or their household members were physically assaulted, had a household member killed, were kidnapped/abducted, were threatened with violence, or experienced sexual violence.

As the core outcomes, our study focused on four key indicators of maternal and child healthcare-seeking. These were i) utilization of antenatal care and ii) facility-based childbirth (delivery in a healthcare facility) for maternal healthcare-seeking, and iii) adherence to the recommended child immunization schedule and iv) use of government health facilities for child healthcare-seeking. Antenatal care use was operationalized as respondents reporting that they received four or more antenatal care visits, the number recommended by the Nigerian Ministry of Health. Facility-based birth is measured as respondents reporting that the child in focus was born in a health clinic or hospital. To measure immunization coverage, interviewers checked the child’s official immunization card to verify whether they had received the full schedule of vaccinations recommended by the Nigerian Ministry of Health. To operationalize the use of government health facilities, we recorded whether individuals used a health facility for any of their children in the past 6 months, and the type of health facility they visited. We then coded reported use of government clinics or hospitals as one, and use of private clinics, traditional healers, and clinics run by religious institutions or other nongovernmental organizations as zero.

A slight uncertainty arises from temporal ordering. In principle, it is possible that some instances of victimization occurred after the health-seeking behavior they are linked to, particularly for maternal indicators, which may refer to events some time before the interview. Although none of the respondents reported such a sequence, we cannot entirely rule out this possibility. Interpretations should therefore be made with greater caution for outcomes that occurred further in the past. This concern does not apply to vaccination status, which was assessed at the time of the interview. Moreover, the usual caveats regarding self-reported behaviors apply, notably that these measures may be affected by imperfect recall or imprecise reporting. As long as such biases affect victimized and non-victimized respondents equally, they would not systematically bias our estimates. However, bias could arise if recall accuracy were directly correlated with victimization status—an effect that cannot be entirely ruled out, though we consider it unlikely. By contrast, measurement of child immunization status is not subject to this concern, as vaccination information was verified from the child’s official immunization card.

### Mediators

In addition to these outcomes, we examined three potential mediating mechanisms: trauma, fear, and mistrust. The hypothesized causal framework is shown in Figure 1. Following prior research in conflict settings, trauma is most often assessed in terms of post-traumatic stress disorder (PTSD) and related symptomatology, typically using standardized instruments such as the Harvard Trauma Questionnaire or the PTSD Checklist (PCL) [60, 61]. PTSD reflects the psychological sequelae of past traumatic events [62], and in our study it was measured using the validated eight-item PTSD Checklist (PCL-8) [63]. Fear, by contrast, captures a forward-looking perception of continued threat, closely linked to the anticipation of future violence rather than past exposure [64]. In previous work, fear has been operationalized through self-reports of avoidance behaviors or through predicted local violence risk in conflict-affected settings [17, 19, 65]. We followed the self-report approach by asking respondents directly about their perceived likelihood to become victims of violence again in the future. The third mediator, institutional mistrust, refers to the erosion of confidence in the state’s ability or willingness to fulfill its protective and service-providing roles [66, 67]. Victimization implies that the state has failed in its role as protector, which may undermine confidence not only in security institutions but also in other state services such as healthcare. This spillover matters for health outcomes: mistrustful individuals are less likely to seek care, follow medical advice, or vaccinate their children [17, 21, 42, 68]. To capture this dimension, our survey included a question on whether respondents’ trust in healthcare workers had increased, decreased, or remained unchanged over the past three years. The full wording of all survey questions can be found in Table S14 in the SI.

**Figure 1:**
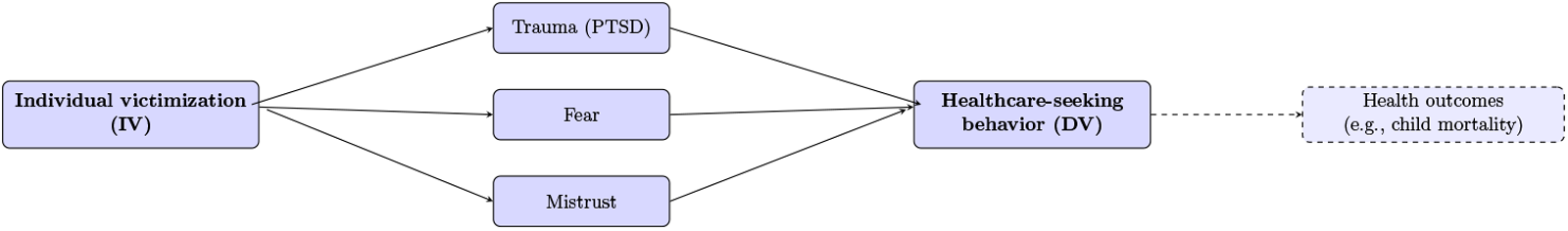
Hypothesized causal framework. *Note*: The figure shows the hypothesized causal relations explored in this paper. Individual victimization is assumed to affect healthcare-seeking behavior through trauma, fear, and mistrust. Arrows reflect temporal/causal ordering. The dashed arrow from healthcare-seeking behavior to health outcomes (e.g., child mortality) indicates the next logical link in the chain but is not estimated here. For an analysis testing the full causal chain with aggregate data, see [17].

To conduct the mediation analysis, we adopt the causal mediation framework proposed by Imai et al. [48]. In practice, we use the unconditional logistic regression outcome model as the basis for the mediation analysis. We estimate separate regression models for each mediator conditional on victimization and covariates, and then decompose the total effect into the natural indirect effect (mediation pathway), the direct effect, and the proportion mediated (PM). To account for the hypothesized presence of multiple mediators, we include the respective other mediators as controls in each model [69]. A core identifying assumption of this approach is sequential unconfoundedness, meaning that there are no unmeasured confounders of the treatment–outcome or mediator–outcome relationships. In practice, this assumption is difficult to fully satisfy, even when controlling for a rich set of covariates as in the present analysis. Therefore, the mediation results should be interpreted with appropriate caution.

Finally, we conducted a complementary regression analysis to identify predictors of declining trust in healthcare workers. The outcome was a binary indicator of whether respondents reported that their trust in healthcare workers had decreased over the past three years. We estimated our standard matched logistic regression models with predictors including reported absenteeism of healthcare workers, exposure to violence by security forces, fear for security at health facilities, worry about undue political influence on the health system, and the provision of free healthcare provided by NGOs. All models adjust for the same sociodemographic covariates used in the main models. All predictors were measured through self-report survey items (full wording in Table S14, SI). This analysis allows us to examine more directly the contextual factors underlying loss of trust, which emerged as the key mediator in our main models.

## Results

### Sample demographics and matching

Table 1 provides summary statistics for our sample, victimization rates, healthcare-seeking behaviors, and other variables reported in this study. The table reports averages after applying sample weights. The corresponding raw frequencies are reported in the text. Statistics for all variables, including covariates, can be found in Table S4 in the SI, and balance statistics, which list values for victimized and non-victimized individuals separately in line with the STROBE [46] statement, are shown in Table S5, SI. As shown in Table 1, of the 3,006 respondents, 71% (n=2,172) were female and 29% (n=834) were male, with an average age of 36 years. Caregivers provided information on 3,006 children, of whom 48% (n=1,448) were female and 52% (n=1,558) male, with a mean age of 2.5 years. Propensity score matching achieved excellent balance, with none of the mean differences statistically significant and all standardized mean differences were below 7% (see Table S5 and Figure S4 in the SI).^1^

**Table 1:** Summary statistics for sample demographics, predictors, outcomes, and mediators.

|  | Mean | SD | Min | Max | Count |
| --- | --- | --- | --- | --- | --- |
| **Sample demographics** |  |  |  |  |  |
| Respondent female | 0.71 | 0.45 | 0.00 | 1.00 | 3,006 |
| Respondent age | 36.19 | 10.52 | 15.00 | 82.00 | 3,006 |
| Child female | 0.48 | 0.50 | 0.00 | 1.00 | 3,006 |
| Child age | 2.49 | 1.30 | 1.00 | 5.00 | 3,006 |
| **Victimization prevalence** |  |  |  |  |  |
| Was/were physically assaulted | 0.11 | 0.31 | 0.00 | 1.00 | 3,006 |
| Had a household member killed | 0.05 | 0.22 | 0.00 | 1.00 | 3,006 |
| Was/were kidnapped/abducted | 0.05 | 0.21 | 0.00 | 1.00 | 3,006 |

<sup>1</sup> Applying a much stricter caliper (0.025) results in a similar number of matched pairs (647 vs. 651), indicating that most pairs are very closely matched.

|  |  |  |  |  |  |
| --- | --- | --- | --- | --- | --- |
| Witnessed someone else being killed | 0.04 | 0.18 | 0.00 | 1.00 | 3,006 |
| Was/were threatened with violence | 0.03 | 0.16 | 0.00 | 1.00 | 3,006 |
| Experienced sexual violence | 0.01 | 0.10 | 0.00 | 1.00 | 3,006 |
| Victimized during armed violence (any) | 0.21 | 0.41 | 0.00 | 1.00 | 3,006 |
| **Healthcare-seeking behavior** |  |  |  |  |  |
| Child fully vaccinated | 0.62 | 0.48 | 0.00 | 1.00 | 3,006 |
| Recommended antenatal care | 0.77 | 0.42 | 0.00 | 1.00 | 3,006 |
| Birth at medical facility | 0.61 | 0.49 | 0.00 | 1.00 | 3,006 |
| Used government clinic for child | 0.80 | 0.40 | 0.00 | 1.00 | 2,196 |
| **Child health outcomes** |  |  |  |  |  |
| Child had health issue when born | 0.03 | 0.16 | 0.00 | 1.00 | 3,006 |
| Child had health issue during last year | 0.24 | 0.43 | 0.00 | 1.00 | 3,006 |
| **Mediators** |  |  |  |  |  |
| Fear of victimization | 0.47 | 0.50 | 0.00 | 1.00 | 3,006 |
| Post-traumatic stress disorder (PTSD) | 0.24 | 0.43 | 0.00 | 1.00 | 3,006 |
| Loss of trust in healthcare workers | 0.19 | 0.39 | 0.00 | 1.00 | 3,006 |
| **Predictors of loss of trust** |  |  |  |  |  |
| State security perpetrator | 0.08 | 0.27 | 0.00 | 1.00 | 3,006 |
| Feared for security at health facility | 0.03 | 0.18 | 0.00 | 1.00 | 3,006 |
| Worried about undue influence | 0.11 | 0.31 | 0.00 | 1.00 | 3,006 |
| Free healthcare provided by NGO | 0.08 | 0.27 | 0.00 | 1.00 | 3,006 |
| Absenteeism of healthcare workers | 0.19 | 0.40 | 0.00 | 1.00 | 3,006 |

*Note:* Summary statistics for the main outcome and treatment variables used in the study. Reduced number of observations in variable “Used government clinic for child” due to question only asked of individuals reporting use of healthcare.

### Victimization prevalence

Table 1 (and Figure S2 in the SI) also shows the prevalence of different types of victimization in our sample. Common forms of victimization include physical assault, experienced by 11.1% (n=338) of respondents, and the killing or kidnapping of a household member, experienced by 4.9% (n=155) and 4.5% (n=129) of respondents, respectively. Most importantly, overall no fewer than 21% of respondents (n=651) reported being victimized by some form of severe violence, underscoring the importance of considering the impact of individual-level victimization on healthcare-seeking behavior, rather than examining only broader exposure to conflict.

### Healthcare-seeking

As for the prevalence of healthcare-seeking for child and maternal health, full childhood immunization coverage was 61% (n=1,563), significantly below the global average of over 80% [70]. Antenatal care was relatively high compared to other Nigerian states, with 77% (n=2,129) of women attending at least four recommended visits during pregnancy [71]. Children were born in a health clinic or hospital in 61% (n=1,559) of cases, slightly below the 70% estimated for births in low- and middle-income countries (LMICs) [72].

Use of government facilities for child healthcare was reported by 80% (n=1,805) of respondents, reflecting the dominance of the public health sector in the country. Other measures included in the table are discussed below.

### Statistical results

Figure 2 shows the results of the matched case-control logistic regression model, which examines the relationship between healthcare-seeking behavior and violent victimization. Descriptive statistics are shown in Figure S3 in the SI, conditional logistic regression results are shown in Figure S5, and full regression outputs are included in Tables S5–S8. As shown in Figure 2, victimization was not significantly associated with whether respondents attended the recommended number of prenatal care visits (Odds ratio (OR): 1.25, 95% CI: 0.93 to 1.69). Contrary to expectations, the likelihood that a child born to victimized caregivers was delivered in a health clinic or hospital was slightly *higher* than among matched non-victimized individuals (OR: 1.30, 95% CI: 1.01 to 1.66). However, this result lacks full robustness across alternative model specifications (see Tables S9–S11 in the SI).

**Figure 2:**
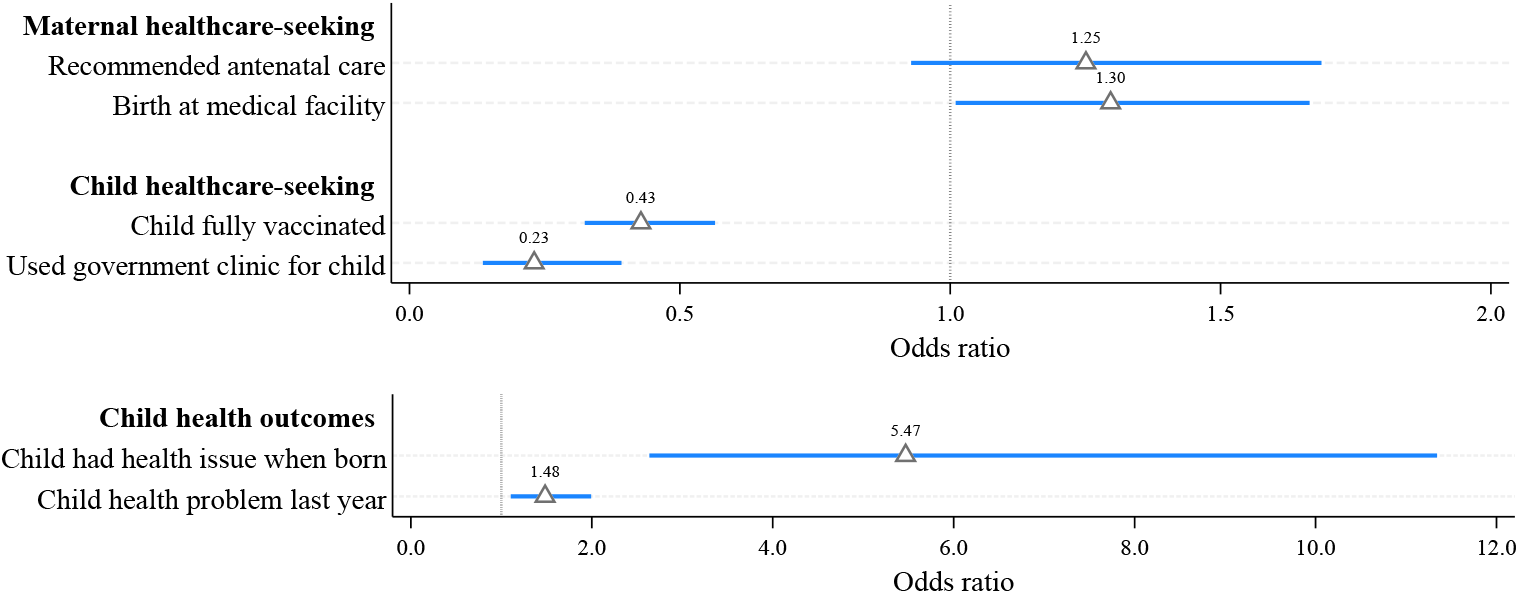
Effect of violent victimization on healthcare-seeking behavior and child health. *Note*: Coefficient plots from a regression of indicators of healthcare-seeking for maternal and child health, and of child health outcomes, on violent victimization. Reported coefficients are odds ratios after unconditional logistic regression using the matched sample. Markers are point estimates, lines 95% confidence intervals. Standard errors clustered at the level of the matched pair.

Thus, healthcare-seeking for maternal health does not differ significantly between victimized and non-victimized individuals. In contrast, there are strong differences in healthcare-seeking for children’s health. Children of victimized caregivers were significantly less likely to be fully vaccinated (OR: 0.43, 95% CI: 0.32 to 0.57). A striking difference emerged in healthcare-seeking *patterns*: children of victimized caregivers were far less likely to be taken to government-run clinics compared to private facilities (OR: 0.23, 95% CI: 0.14 to 0.39).

Figure 2 also shows that victimization was associated with poorer child health. Children of caregivers who experienced violence were less likely to be fully healthy at birth (OR: 5.47, 95% CI: 2.64 to 11.34) and more likely to have recently suffered from health issues such as respiratory, digestion, or coordination problems; or hearing and vision problems (OR: 1.48, 95% CI: 1.10 to 1.99).

To confirm the validity of our findings, we conducted a number of robustness checks. First, conditional logistic regression yields very similar estimates to our main unconditional models: antenatal care (OR: 1.14, 95% CI: 0.89 to 1.45 vs. OR: 1.08), skilled birth attendance (OR: 1.26, 95% CI: 1.01 to 1.59 vs. OR: 1.18), full child vaccination (OR: 0.50, 95% CI: 0.40–0.63 vs. OR: 0.43), and use of government facilities for child care (OR: 0.17, 95% CI: 0.10–0.28 vs. OR: 0.23). Second, we restricted the analysis to female respondents and found that the results remained largely unchanged compared to using the full sample (Table S9). Third, applying small-area (500*×*500m) fixed effects likewise yielded very similar estimates (Table S10, SI). This suggests that our findings are robust even when comparing closely matched individuals living in the same local area, where healthcare supply factors should be constant, making it highly unlikely that the observed differences in healthcare-seeking behavior are driven by variation in service availability rather than the effects of victimization. Finally, probing for differential effects within our main independent variable, we found that the observed negative association between victimization and healthcare-seeking appears to be driven by direct forms of victimization (physical assault, abduction, or sexual violence), whereas indirect victimization (seeing someone killed or losing a household member) did not consistently reduce healthcare-seeking behavior (Table S12, SI).

### Mediation analysis

What explains the lower healthcare-seeking for child health and the shift away from government healthcare among victimized respondents? To investigate this, we examined three key factors discussed in the literature: i) fear of future victimization, which may lead to avoidance behavior; ii) poor mental health, which may impair individuals’ ability to engage in healthcare-seeking behavior; and iii) loss of trust in healthcare personnel, which may prevent victimized individuals from engaging with government institutions.

Table 2 indicates the extent to which the effect of violent victimization on child health is mediated by these three factors, keeping in mind the methodological limitations of this kind of analysis. For childhood vaccination rates, neither fear of future victimization (Proportion mediated (PM): 0.01, 95% CI: −0.09 to 0.12) nor PTSD (PM: 0.01, 95% CI: −0.11 to 0.13) seem to mediate the effect of victimization, despite prior work documenting or suggesting a mediating role for trauma [73–78]. This divergence may reflect characteristics of our setting and measurement: PTSD symptoms were captured with the PCL-8, which may underrepresent avoidance and numbing domains most relevant to preventive care, while contextual constraints such as staff absenteeism, politicization of services, and indeed fear and distrust, may act more proximally on care-seeking behavior than trauma [17, 19]. The non-detection of a mediating relationship does not imply that victimization has no effect on PTSD, however. On the contrary, all three mediators—including fear and PTSD—are strongly predicted by violent victimization, as shown in Table S14 in the SI.

**Table 2:** Mediation analyses.

| Dependent variable: | Child fully vaccinated |  |  | Used government clinic for child |  |  |
| --- | --- | --- | --- | --- | --- | --- |
|  | (1)<br>FearVict | (2)<br>PTSD | (3)<br>DistrustMed | (4)<br>FearVict | (5)<br>PTSD | (6)<br>DistrustMed |
| Nat. indirect effect | -0.001<br>(-0.011 to 0.009) | -0.001<br>(-0.010 to 0.009) | -0.041**<br>(-0.073 to -0.009) | -0.030***<br>(-0.050 to -0.010) | 0.003<br>(-0.002 to 0.008) | -0.089***<br>(-0.137 to -0.041) |
| Total effect | -0.096***<br>(-0.150 to -0.042) | -0.082***<br>(-0.138 to -0.026) | -0.128***<br>(-0.174 to -0.082) | -0.073**<br>(-0.138 to -0.008) | -0.052*<br>(-0.110 to 0.006) | -0.127***<br>(-0.173 to -0.080) |
| Proportion mediated (PM) | 0.01<br>(-0.09 to 0.12) | 0.01<br>(-0.11 to 0.13) | 0.32**<br>(0.03 to 0.61) | 0.41*<br>(-0.04 to 0.86) | -0.06<br>(-0.17 to 0.06) | 0.70***<br>(0.25 to 1.16) |
| N | 1,302 | 1,302 | 1,302 | 979 | 979 | 979 |

*Note:* Mediation analysis for the effect of violent victimization on child healthcare-seeking. Models 1–3 show mediation analyses for our indicator as to whether a child is fully vaccinated, Models 4–6 show mediation analyses for the effect of victimization on child healthcare-seeking at government health clinics or hospitals. Nat. indirect effect stands for natural indirect (mediated) effect. FearVict records respondents reporting fear of future violence, PTSD measures respondents suffering from post-traumatic stress disorder, and DistrustMed records whether respondents report a loss of trust in healthcare workers. Reported coefficients are marginal effects after logistic regression. Matched sample of victimized and non-victimized respondents. Proportions mediated do not sum to 100 since baseline values vary (the total effect is estimated conditional on the other mediators). Standard errors clustered at the level of the matched pair, 95% confidence intervals in parentheses, \* $p < 0.1$ , \*\* $p < 0.05$ , \*\*\* $p < 0.01$ .

Supporting this idea, loss of trust in healthcare workers emerges as a strong and statistically significant mediator, explaining approximately one third (PM: 0.32, 95% CI: 0.03 to 0.61) of the effect of victimization. The mediation pattern is more complex for children’s healthcare-seeking in government facilities. Here, 41% of the effect of victimization (controlling for other mediators) could be attributed to perceived insecurity, a result that approaches conventional statistical significance (PM: 0.41, 95% CI: −0.04 to 0.86). PTSD, once again, does not mediate the effect of victimization (PM: −0.06, 95% CI: −0.17 to 0.06). However, mistrust in healthcare providers appears to have an extraordinarily strong mediating effect, accounting for no less than 70% of the conditional effect of victimization (PM: 0.70, 95% CI: 0.25 to 1.16). Our analysis thus strongly suggests that fear and especially distrust play important mediating roles. While these results align with previous research, they provide more direct empirical evidence linking both factors to victimization rather than to general exposure to armed violence. In contrast, despite its theoretical plausibility and prior empirical support, in our setting poor mental health (PTSD) does not appear to explain the observed association between victimization and healthcare-seeking for children.

### Determinants of the loss of trust in healthcare workers

Given the important role of a loss of trust in mediating healthcare-seeking behavior, we investigated its underlying determinants in the final step of our analysis. A key finding was that loss of trust in health workers was strongly correlated with loss of trust in other state institutions. Specifically, individuals who lose trust in the government (OR: 35.76, 95% CI: 23.72 to 53.91) and in the security forces (OR: 23.76, 95% CI: 17.43 to 32.38) were significantly more likely to lose trust in healthcare providers. These findings suggest that distrust in healthcare workers extends beyond the health sector, reflecting a broader erosion of trust in state institutions. This generalized distrust may result from a perceived failure of the government or security forces to protect individuals from violence, leading victims to associate the healthcare system with the same institutions they hold responsible for their victimization. Second, we regress the indicator for loss of trust in healthcare workers on several conflict-related experiences reported in our survey. Specifically, we focus on whether distrust is linked to: i) violent victimization at the hands of state security forces, ii) healthcare worker absenteeism when respondents sought medical assistance, iii) fear for personal safety while at a health facility, iv) concerns that health facilities were unduly influenced by the government, or v) receiving free healthcare from nongovernmental organizations (NGOs).

Victimization by state security forces is alarmingly common in our sample. Among those who experienced violence, more than one-third of victimized respondents (38%) – equivalent to 8% (n=212) of the full sample, see Table 1 – reported that state security forces were the perpetrators. Such experiences are likely to undermine trust in the government, which in turn may spill over to health workers [66, 79, 80].

Similarly, health worker absenteeism, experienced by 17% (n=527) of respondents, has previously been found to correlate with lower levels of trust [81]. Concerns about the security of health facilities, cited by 4% (n=122) of respondents, and about undue political influence on health facilities, cited by 13% (n=396) of respondents, may also contribute to mistrust. In contrast, receiving free healthcare from NGOs, reported by 9% (n=272) of respondents, may have the opposite effect. Previous research suggests that humanitarian interventions providing free medical care can mitigate the negative effects of conflict exposure [18], and it is plausible that access to such services helps to restore trust in healthcare workers [68].

Figure 3 shows the results of the regression analysis. The dominant factors explaining the loss of trust in healthcare are victimization by state security forces (OR: 30.23, 95% CI: 17.64 to 51.81) and absenteeism of healthcare workers (OR: 13.43, 95% CI: 9.58 to 18.85). Other significant, but less pronounced, predictors include: feeling unsafe when seeking healthcare (OR: 2.01, 95% CI: 1.10 to 3.70) and worrying about undue political influence on the healthcare system (OR: 1.50, 95% CI: 1.02 to 2.20). Meanwhile, receiving free healthcare is associated with a relative gain in trust, but not significantly so (OR: 0.76, 95% CI: 0.47 to 1.22). Overall, the results clearly show how failed attempts to access healthcare and negative experiences with other government institutions contribute to a decline in trust.

**Figure 3:**
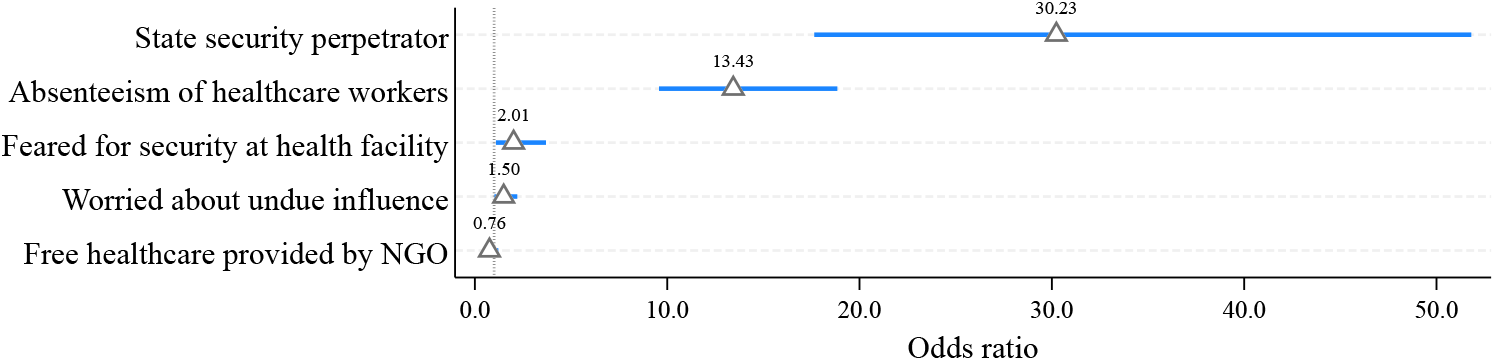
Determinants of loss of trust in healthcare workers. *Note*: Coefficient plots from a regression of the indicator for loss of trust in healthcare workers on various potential explanatory variables. Reported coefficients are odds ratios after unconditional logistic regression using the matched sample. Markers are point estimates, lines 95% confidence intervals. Standard errors clustered at the level of the matched pair.

## Conclusion

Our study advances the literature on the health effects of armed conflict in three key ways. First, we focus explicitly on the effects of individual-level victimization, a quantitatively significant yet often overlooked consequence of armed conflict. Unlike broad measures of exposure to conflict, which are often defined in terms of proximity to violent events, victimization is a more direct and specific experience. In our sample, 21% of respondents reported experiencing severe forms of victimization, such as being violently attacked or having a household member killed. These respondents exhibit markedly different healthcare-seeking behaviors compared to non-victimized (but still conflict-exposed) individuals living in close proximity, underscoring the importance of looking beyond general exposure. Second, echoing other recent research [9, 15–19, 22], we focus on demand-side effects, i.e., healthcare-seeking, rather than on supply-side factors such as the destruction of infrastructure. By adjusting for healthcare supply factors, such as proximity to health facilities or availability of health workers, and by using small-area fixed effects, we effectively isolated the effect of violence on the demand for healthcare. We found that while maternal health-seeking was resilient to victimization, child health outcomes showed acute vulnerability, with children of victimized caregivers having significantly lower vaccination rates and much lower rates of presentation to government health facilities. This is consistent with other studies that have found that pediatric care tends to be neglected in conflict-affected areas [18, 82]. Third, looking beyond top-level associations, we present evidence for a mechanism by which individual-level victimization translates into lower healthcare-seeking for child health and a shift in healthcare-seeking away from government-run facilities: a loss of trust in healthcare workers. We found that the loss of trust was most strongly associated with absenteeism among healthcare workers and instances of violent victimization by state security forces. Contrary to theoretical expectations [36, 37], victimization-induced trauma did not mediate the relationship between victimization and healthcare-seeking behavior.

As always, this study is not without limitations. For one, the cross-sectional nature of the data limits our ability to definitively establish causality. Although the matched case-control design strengthens causal inference, we cannot completely rule out the possibility that unobserved confounders may have influenced the results. In addition, the study focuses on a specific region in Nigeria, which may limit the generalizability of the findings to other conflict-affected contexts. Future research should examine these relationships in different settings and strengthen causal inference by using longitudinal data to track causal pathways linking victimization, trust, and health-seeking behaviors over time.

While more research is needed to validate the results, our findings suggest several policy priorities. A first area of intervention is to prevent the loss of trust. Our findings suggest that to achieve this goal, at least basic emergency services should be provided throughout a violent conflict, and security forces should exercise the utmost discipline and care to avoid harming civilians. A second area of intervention is to mitigate the consequences where trust has already been lost and healthcare-seeking for children has been reduced. While other work has found that aid provided by NGOs can help counteract the negative effects of exposure to armed conflict on trust [18, 68], our evidence suggests that such aid may not be sufficient to rebuild trust among actual victims.

This calls for a deeper exploration of how trust might be rebuilt. One possible strategy is targeted child health interventions, for instance in the form of community engagement strategies [83–85], which could help rebuild state-community relations. Political science research also suggests that legal accountability for perpetrators is important for rebuilding trust [86], and that listening exercises that explicitly acknowledge suffering can be used to restore trust in the state after conflict [87–89]. Such exercises could take the form of outreach and communication campaigns by health authorities—for example, acknowledging hardships and encouraging families to return to government health services. Combining such symbolic acknowledgment with visible improvements in service quality may be particularly effective, as it couples recognition with tangible action and incentives [90].

Evidence from other conflict-affected settings shows that interventions can be designed to both improve health outcomes and rebuild trust in service delivery. Community Health Worker (CHW) programs, which relied on locally recruited workers and participatory planning, improved immunization coverage and strengthened community confidence in the health system [91–93]. Similar approaches could be adapted in Nigeria, where trust has likewise proven central to sustained healthcare utilization [94]. Experience from Northern Nigeria and elsewhere demonstrate that adaptive, multi-level engagement with governance, financing, and community systems can strengthen health system resilience even under insecurity [95, 96]. Building on such principles, displaced persons or youth volunteers could be trained as health promoters in IDP camps and host communities, and mobile units staffed by female health workers could enhance access and trust among women in conservative or insecure areas [97, 98]. Conflict-sensitive vaccination campaigns that engage local and religious leaders to counter misinformation, as successfully implemented elsewhere [99, 100], could likewise inform strategies to overcome vaccine hesitancy and insecurity. These approaches must, however, be carefully tailored to Nigeria’s sociopolitical realities, including collaboration with religious institutions and strict avoidance of coercive associations with security actors.

Ultimately, our findings underscore that in contexts of protracted conflict, rebuilding trust is as central to improving health outcomes as restoring supply, and future interventions should be rigorously evaluated with this dual objective in mind.

## Contributorship

MS, GCA, and LA developed the research idea. All co-authors helped to develop the research design and survey instrument. RIJ and MS supervised the fieldwork, and AAA, AAK, MIJ, and AM led the interview data collection. MA and LK performed statistical analyses and interpreted results. MS drafted the initial manuscript, with extensive input from LA. All members reviewed and provided critical feedback. MS serves as guarantor.

## Ethics

Interviews were conducted by trained interviewers in the local language (Hausa) using a pre-tested questionnaire. Participants were informed in advance of the sensitive nature of some of the questions included in the survey, and were informed that they could end the survey or skip individual questions at any time. Only individuals who gave their explicit informed consent were interviewed. The survey and data collection protocol were approved by the National Health Research Ethics Committee of Nigeria (NHREC; Approval No. NHREC/01/01/2007-14/01/2024), by the respective authorities of the three states, and by the University of Konstanz, Germany (Institutional Review Board (IRB) Statement 13/2024).

## Data availability statement

Data are available in a public, open access repository. The de-identified survey microdata, replication code, and accompanying materials are available on the Open Science Framework (OSF) at: https://doi.org/10.17605/OSF.IO/6DE2R. Please cite the dataset as: [dataset] [47] Schaub M, Adeyanju GC, Abreu L. Replication data for: Maternal and child health-seeking among victims of violence in armed conflict: A matched case-control study in northeast Nigeria. Open Science Framework (OSF), February 16, 2025. https://doi.org/10.17605/OSF.IO/6DE2R.

## Footnotes

1 Applying a much stricter caliper (0.025) results in a similar number of matched pairs (647 vs. 651), indicating that most pairs are very closely matched.

