## Supplementary material for "Maternal and child healthcare-seeking among victims of violence in armed conflict: A quasi-experimental study in northeast Nigeria∗": SI

#### Table of Contents


| | | |
|---|---|---|
| **A** | **Consensus statement** | **1** |
| **B** | **STROBE reporting checklist** | **3** |
| **C** | **Sampling area, sample size and participation rates** | **4** |
| **D** | **Summary statistics** | **6** |
| **E** | **Descriptive outcomes** | **7** |
| **F** | **Balance statistics** | **9** |
| **G** | **Regression output** | **10** |
| **H** | **Conditional logistic regression** | **11** |
| **I** | **Consequences for child health** | **12** |
| **J** | **Robustness checks** | **13** |
| **K** | **Direct effect on mediators** | **17** |
| **L** | **Survey question wording** | **18** |

### A Consensus statement

Table S1: Consensus statement

| Category/Question | Response |
|---|---|
| **Study Conceptualisation** | |
| How does this study address local research and policy priorities? | Our study examines maternal and child health in three states in northeastern Nigeria.This was already an area of active research for all the Nigerian institutions involved in the study, and is therefore closely aligned with local priorities.The focus on the health impact of armed violence is particularly important because of the high burden that the armed insurgency has placed on local populations and health workers, and the fact that health outcomes have deteriorated in the wake of the insurgency. A better understanding of the reasons for this is crucial to improving service delivery and health outcomes. |
| How were local researchers involved in study design? | Local researchers participated in the conceptualization and design. The VIVAC team held its first meeting at Aminu Kano Teaching Hospital, Nigeria, in February 2023.The research idea was fully discussed and inputs were provided by all members, including national and international collaborators. |
| **Research Management** | |
| How has funding been used to support the local research team(s)? | Funding supported training in research ethics, research methods, and grant writing, with some courses offered online. |
| **Data Acquisition and Analysis** | |
| How are research staff who conducted data collection acknowledged? | State enumerators, supervised by state coordinators, generated the data. Their contributions earned them authorship, and all contributors beyond the authors were acknowledged. |
| How have members of the research partnership been provided with access to study data? | Partners accessed study data through a cloud-based platform with secure login credentials per state and for the Nigeria Project Lead. |
| How were data used to develop analytical skills within the partnership? | Shared data were reviewed and discussed both formally and informally, including communication between state coordinators and the national team leader for clarifications. |
| **Data Interpretation** | |
| How have research partners collaborated in interpreting study data? | All results were interpreted with state teams to ensure local relevance. Findings were discussed in bi-weekly research team meetings. |
| **Drafting and Revising for Intellectual Content** | |
| How were research partners supported to develop writing skills? | Partners participated in report and manuscript writing. After fieldwork, team members documented activities, lessons learned, and challenges to guide future research. |
| How will research products be shared to address local needs? | Findings will be shared with local stakeholders on an ongoing basis, disseminated through meetings in all states, and published for easy access by the public. A first dissemination meeting was held in January 2025 with representatives of the maternal and child health services in the Ministries of Health of Adamawa, Borno and Yobe states, the WHO Nigeria Country Office and the National Primary Healthcare Development Agency (NPHCDA). |
| **Authorship** | |
| How is the leadership, contribution, and ownership of this work by LMIC researchers recognized? | Leadership and ownership were established through appropriate authorship positions. More than half (7/11) of co-authors are LMIC researchers. |

*Continued on next page*

*Continued from previous page*

| **Category/Question** | **Response** |
|---|---|
| How have early career researchers across the partnership been included?<br>How has gender balance been addressed within the authorship? | At least 3 national collaborators are early career researchers with fewer than 10 publications. They were given priority in authorship. The team is gender-balanced, with 5 of 11 authors being female, ensuring diversity, equity, and inclusion. |
| **Training** | |
| How has the project contributed to training of LMIC researchers? | Five LMIC co-authors received training in health research ethics, research methods, and data analysis at the start of the project. Enumerators were trained in interviewing methods – skills that will be transferable to future work. |
| **Infrastructure** | |
| How has the project contributed to improvements in local infrastructure? | The project provided 24 data collection tablets and 6 digital audio recorders to Bayero University & Aminu Kano Teaching Hospital, Nigeria. |
| **Governance** | |
| What safeguarding procedures were used to protect local study participants and researchers? | Participants were interviewed in safe environments. Security personnel were employed in high-risk areas, ensuring researchers' safety. |

### B STROBE reporting checklist

*Note*: The study follows the STROBE guidelines for observational research [1] and adapts selected items to a quasi-experimental, propensity-score–matched design.

Table S2: STROBE reporting

| **Reporting Item** | **Item Nr.** | **Reporting Item** | **Page Nr.** |
|---|---|---|---|
| Title | #1a | Indicate the study's design with a commonly used term in the title or the abstract | 1 |
| Abstract | #1b | Provide in the abstract an informative and balanced summary of what was done and what was found | 1 |
| Background / rationale | #2 | Explain the scientific background and rationale for the investigation being reported | 4 |
| Objectives | #3 | State specific objectives, including any prespecified hypotheses | 4 |
| Study design | #4 | Present key elements of study design early in the paper | 5 |
| Setting | #5 | Describe the setting, locations, and relevant dates, including periods of recruitment, exposure, follow-up, and data collection | 5 |
| Eligibility criteria | #6a | Give the eligibility criteria, and the sources and methods of study inclusion. | 5 |
| Eligibility criteria | #6b | For matched studies, give matching criteria and the number of controls per treated case | 6 |
| | #7 | Clearly define all outcomes, exposures, predictors, potential confounders, and effect modifiers. Give diagnostic criteria, if applicable | 6–7 |
| Data sources / measurement | #8 | For each variable of interest give sources of data and details of methods of assessment (measurement) | 6–7, SI 15–19 |
| Bias | #9 | Describe any efforts to address potential sources of bias | 9 |
| Study size | #10 | Explain how the study size was arrived at | 5, SI 4 |
| Quantitative variables | #11 | Explain how quantitative variables were handled in the analyses | 8, SI 15–19 |
| Statistical methods | #12a | Describe all statistical methods, including those used to control for confounding | 6, 9 |
| Statistical methods | #12b | Describe any methods used to examine subgroups and interactions | SI 11 |
| Statistical methods | #12c | Explain how missing data were addressed | SI 5 |
| Statistical methods | #12d | If applicable, explain how matching of cases and controls was addressed | 6 |
| Statistical methods | #12e | Describe any sensitivity analyses | SI 11–13 |
| Participants | #13a | Report numbers of individuals at each stage of study | SI 4 |
| Participants | #13b | Give reasons for non-participation at each stage | SI 4 |
| Participants | #13c | Consider use of a flow diagram | SI 4 |
| Descriptive data | #14a | Give characteristics of study participants | 8, SI 5 |
| Descriptive data | #14b | Indicate number of participants with missing data for each variable of interest | SI 5 |
| Outcome data | #15 | Report numbers in each exposure category | SI 7 |
| Main results | #16a | Give unadjusted estimates and confounder-adjusted estimates | SI 13 |
| Main results | #16b | Report category boundaries when continuous variables were categorized | NA |
| Main results | #16c | Consider translating estimates of relative risk into absolute risk | NA |
| Other analyses | #17 | Report other analyses done | 10, SI 11–13 |
| Key results | #18 | Summarise key results with reference to study objectives | 11–12 |
| Limitations | #19 | Discuss limitations of the study, considering bias or imprecision | 12 |
| Interpretation | #20 | Give a cautious overall interpretation | 12 |
| Generalisability | #21 | Discuss the generalisability of the study results | 12 |
| Funding | #22 | Give the source of funding and the role of the funders | 3 |

### C Sampling area, sample size and participation rates

#### Sampling area

Figure S1: Overview of study area

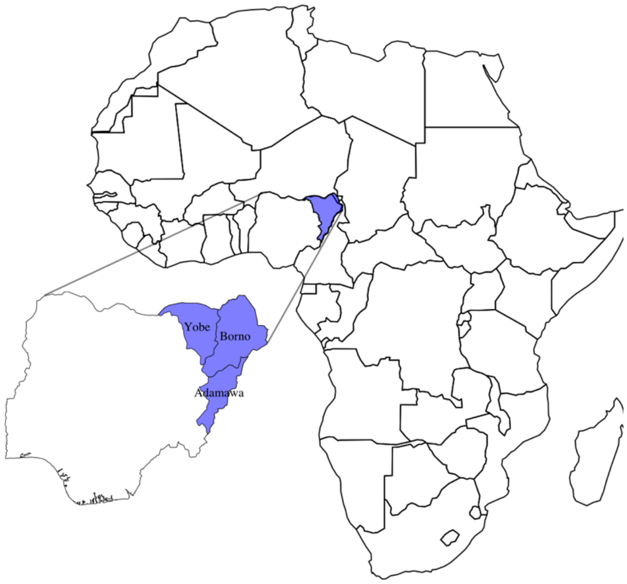


*Note:* The figure shows the northeastern Nigerian states of Adamawa, Borno, and Yobe in the regional context.

#### Sample size considerations

For estimating the prospective sample size, we used the share of fully vaccinated children as the target outcome since we knew that we would be able to record this outcome with high fidelity (by reviewing immunization cards). Sampling size was based on the assumption that this share would be 60% in the non-victimized population, a figure derived from the Nigerian team's previous experience with smaller case numbers. Our goal was to detect a 10% point lower vaccination rate among victimized individuals with 0.8 power and 95% confidence. To do this, we would need 388 individual cases (i.e., individuals who were victims of armed violence). Assuming the rate of victimization to be 20%, we therefore would need to recruit a minimum of 388/.2=1,940 individuals. A further consideration was that this sample size would also allow the use of four times the number of treated units from which to select matched controls, a ratio well in line with the relevant literature [2]. As these numbers are highly sensitive to small deviations from the assumptions, we decided to aim for a sample size of n=3,000, i.e., a sample size that is 1,000 observations larger.

##### Participation rates

We attempted to recruit a total of 3,438 individuals. In 432 cases, recruitment attempts were unsuccessful or observations had to be excluded for quality reasons, for a total sample size of 3,006. The breakdown of the reasons for exclusion is provided in Table S3.

Table S3: Survey participation rates

| | |
|---|---|
| Total contact attempts made | 3,438 |
| Total unsuccessful/excluded | 432 |
| *Reason for exclusion** | |
| No one at home | 109 |
| Refused | 12 |
| Incomplete | 121 |
| Too short | 155 |
| Incorrect location | 158 |
| Complete observations | 3,006 |
| Participation rate | 87% |

*Numbers do not add up to "Total unsuccessful/excluded" because several reasons can apply to the same observation.

### D Summary statistics

Table S4: Summary statistics, full table

| | Mean | SD | Min | Max | Count |
|---|---|---|---|---|---|
| **Variables in focus** | | | | | |
| Victimized during armed violence | 0.21 | 0.41 | 0.00 | 1.00 | 3,006 |
| Child fully vaccinated | 0.62 | 0.48 | 0.00 | 1.00 | 3,006 |
| Recommended antenatal care | 0.77 | 0.42 | 0.00 | 1.00 | 3,006 |
| Birth at medical facility | 0.61 | 0.49 | 0.00 | 1.00 | 3,006 |
| Used government clinic for child | 0.80 | 0.40 | 0.00 | 1.00 | 2,196 |
| Child had health issue when born | 0.03 | 0.16 | 0.00 | 1.00 | 3,006 |
| Child had health issue during last year | 0.24 | 0.43 | 0.00 | 1.00 | 3,006 |
| PTSD according to PCL8-5 index | 0.24 | 0.43 | 0.00 | 1.00 | 3,006 |
| Loss of trust in healthcare workers | 0.19 | 0.39 | 0.00 | 1.00 | 3,006 |
| State security perpetrator | 0.08 | 0.27 | 0.00 | 1.00 | 3,006 |
| Feared for security at health facility | 0.03 | 0.18 | 0.00 | 1.00 | 3,006 |
| Worried about undue influence | 0.11 | 0.31 | 0.00 | 1.00 | 3,006 |
| Free healthcare provided by NGO | 0.08 | 0.27 | 0.00 | 1.00 | 3,006 |
| Absenteeism of healthcare workers | 0.19 | 0.40 | 0.00 | 1.00 | 3,006 |
| **Control variables** | | | | | |
| Female | 0.71 | 0.45 | 0.00 | 1.00 | 3,006 |
| Age | 36.19 | 10.52 | 15.00 | 82.00 | 3,006 |
| Education | 2.67 | 1.53 | 1.00 | 8.00 | 3,006 |
| Number of children | 5.18 | 3.86 | 1.00 | 60.00 | 3,006 |
| Child female | 0.48 | 0.50 | 0.00 | 1.00 | 3,006 |
| Age of child | 2.49 | 1.30 | 1.00 | 5.00 | 3,006 |
| Muslim | 0.76 | 0.42 | 0.00 | 1.00 | 3,006 |
| Walking time to closest health facility | 32.54 | 32.86 | 0.00 | 280.00 | 3,006 |
| N healthcare facilities | 0.20 | 0.45 | 0.00 | 2.00 | 3,006 |
| DPT-3 coverage | 0.29 | 0.15 | 0.05 | 0.44 | 3,006 |
| Measles vaccination coverage | 54.23 | 6.19 | 29.68 | 63.86 | 3,006 |
| Under-5 mortality | 19.42 | 1.59 | 16.48 | 23.47 | 3,006 |
| Underweight prevalence | 25.63 | 6.98 | 16.09 | 40.59 | 3,006 |
| Malaria prevalence, age 0-4 | 0.34 | 0.21 | 0.19 | 1.31 | 3,006 |
| Female education, age 20-24 | 4.95 | 1.20 | 1.66 | 6.85 | 3,006 |
| Population density | 59.94 | 52.66 | 0.00 | 190.20 | 3,006 |
| Distance to major road | 2.24 | 4.66 | 0.01 | 28.22 | 3,006 |
| Road distance to regional capital | 2.03 | 1.60 | 0.00 | 5.37 | 3,006 |

*Note:* Summary statistics for variables used in the study. Individual-level variables recoded to binary from survey questions reproduced in Table S14 below. Missing values coded as zero. Reduced number of observations in variable "Used government clinic for child" due to question only asked of individuals reporting use of health care.

### E Descriptive outcomes

Figure S2: Prevalence and type of violent victimization

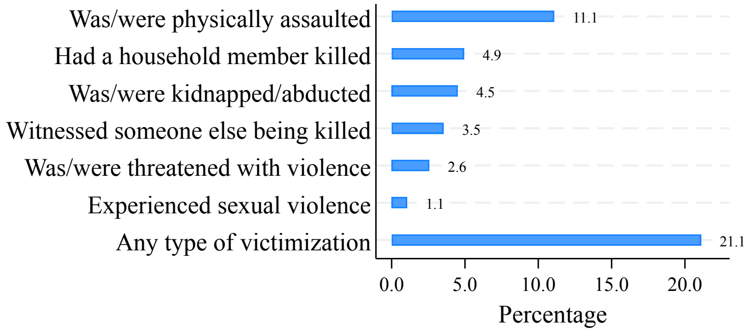


*Note:* The figure shows the prevalence and types of violent victimization suffered by respondents at the hands of armed actors, n=3,006. The data are representative of Adamawa, Borno, and Yobe states, Nigeria.

Figure S3: Descriptive differences in healthcare-seeking

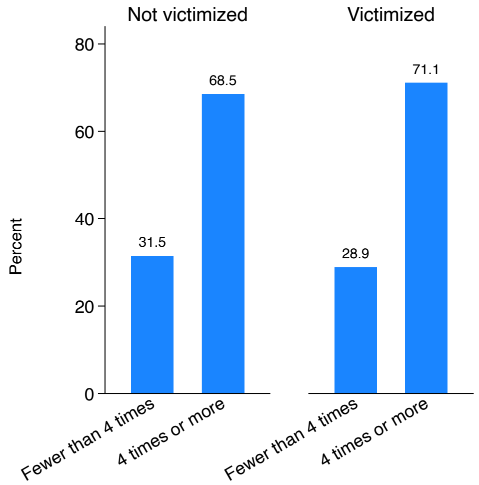


(a) Recommended antenatal care

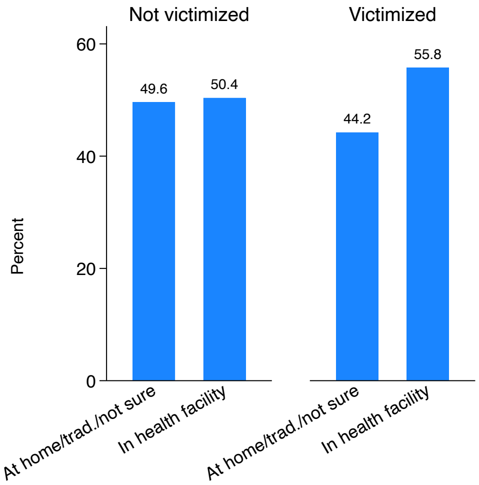


(b) Birth at medical facility

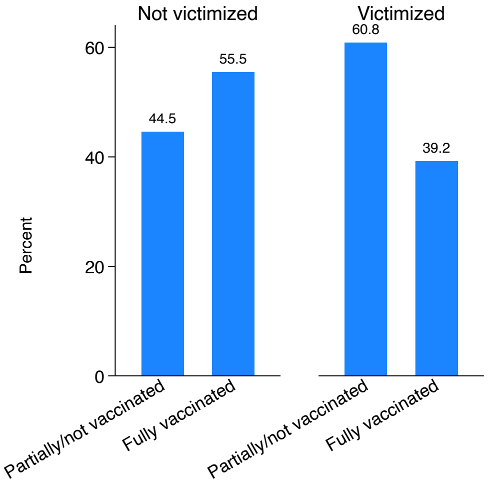


(c) Child fully vaccinated

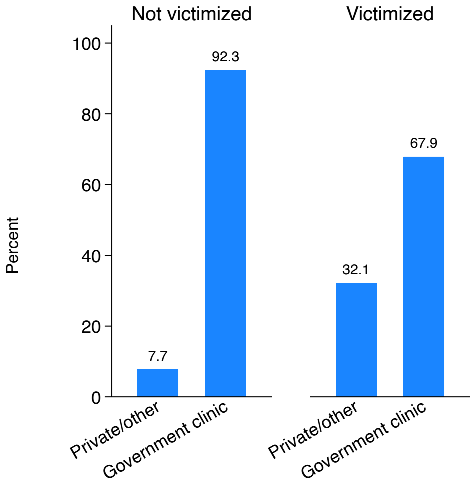


(d) Used government clinic for child

*Note:* The figure shows the descriptive differences in healthcare-seeking between victimized and non-victimized respondents in the matched sample with 651 cases and 651 controls.

### F Balance statistics

Table S5: Balance after matching

| | Not victimized | | | Victimized | | | |
|---|---|---|---|---|---|---|---|
| | N | Mean | SD | N | Mean | SD | Diff |
| Female | 651 | 0.762 | 0.426 | 651 | 0.774 | 0.418 | 0.012 |
| Age | 651 | 34.207 | 10.123 | 651 | 33.955 | 9.511 | -0.252 |
| Education | 651 | 2.367 | 1.506 | 651 | 2.333 | 1.555 | -0.034 |
| Number of children | 651 | 4.416 | 2.946 | 651 | 4.206 | 2.961 | -0.210 |
| Child female | 651 | 0.452 | 0.498 | 651 | 0.464 | 0.499 | 0.012 |
| Age of child | 651 | 2.307 | 1.242 | 651 | 2.255 | 1.331 | -0.052 |
| Muslim | 651 | 0.720 | 0.449 | 651 | 0.737 | 0.440 | 0.017 |
| Walking time to closest health facility | 651 | 39.564 | 36.560 | 651 | 38.296 | 37.501 | -1.267 |
| N healthcare facilities | 651 | 0.124 | 0.374 | 651 | 0.124 | 0.340 | 0.000 |
| DPT-3 coverage | 651 | 0.238 | 0.131 | 651 | 0.232 | 0.130 | -0.006 |
| Measles vaccination coverage | 651 | 50.164 | 10.939 | 651 | 50.047 | 10.799 | -0.116 |
| Under-5 mortality | 651 | 20.505 | 1.498 | 651 | 20.569 | 1.493 | 0.064 |
| Underweight prevalence | 651 | 28.156 | 4.852 | 651 | 28.488 | 4.865 | 0.332 |
| Malaria prevalence, age 0-4 | 651 | 0.405 | 0.246 | 651 | 0.403 | 0.242 | -0.003 |
| Female education, age 20-24 | 651 | 4.139 | 1.257 | 651 | 4.075 | 1.245 | -0.064 |
| Population density | 651 | 28.657 | 34.462 | 651 | 29.387 | 36.097 | 0.731 |
| Distance to major road | 651 | 3.417 | 5.812 | 651 | 3.205 | 5.686 | -0.212 |
| Road distance to regional capital | 651 | 2.545 | 1.513 | 651 | 2.468 | 1.463 | -0.077 |

* $p < 0.1$, ** $p < 0.05$, *** $p < 0.01$. *Note:* No statistically significant differences in Table.

Figure S4: Standardized mean differences between cases and controls

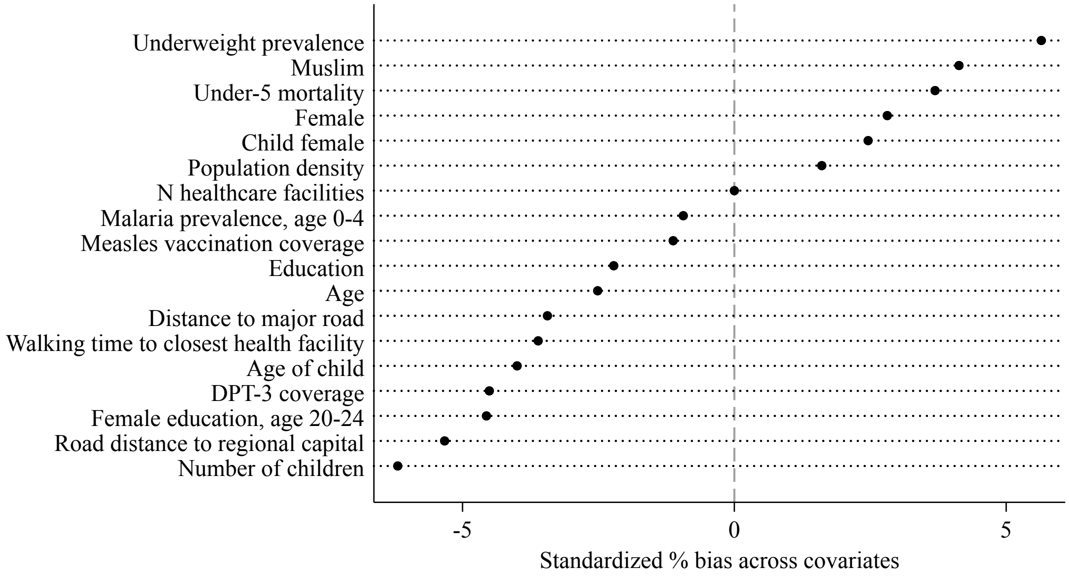


*Note:* The figure shows the remaining standardized mean bias between victimized and non-victimized individuals after matching.

### G Regression output

Table S6: Effect of violent victimization on healthcare-seeking behavior

| | (1) AntenRec | (2) BirthMed | (3) ChildVacc | (4) ChildGov |
|---|---|---|---|---|
| Victimized during armed violence | 1.25 | 1.30** | 0.43*** | 0.23*** |
| | [0.93,1.69] | [1.01,1.66] | [0.32,0.57] | [0.14,0.39] |
| Female | 1.75*** | 1.27 | 1.97*** | 0.38** |
| | [1.17,2.62] | [0.89,1.81] | [1.33,2.91] | [0.17,0.83] |
| Age | 1.01 | 1.02** | 1.02* | 0.98 |
| | [0.99,1.03] | [1.00,1.04] | [1.00,1.03] | [0.95,1.02] |
| Education | 1.05 | 1.12** | 1.21*** | 1.32*** |
| | [0.93,1.19] | [1.03,1.22] | [1.10,1.33] | [1.14,1.53] |
| Number of children | 0.92** | 0.92*** | 1.00 | 1.27*** |
| | [0.87,0.98] | [0.88,0.97] | [0.95,1.05] | [1.07,1.50] |
| Child female | 1.28 | 1.01 | 0.85 | 1.03 |
| | [0.94,1.74] | [0.78,1.31] | [0.64,1.12] | [0.67,1.58] |
| Age of child | 0.90 | 1.01 | 2.04*** | 1.31** |
| | [0.79,1.02] | [0.91,1.12] | [1.77,2.37] | [1.05,1.65] |
| Muslim | 0.53** | 1.19 | 0.38*** | 0.19*** |
| | [0.32,0.90] | [0.76,1.86] | [0.24,0.61] | [0.07,0.53] |
| Walking time to closest health facility | 0.99*** | 1.00 | 1.01*** | 0.98*** |
| | [0.98,1.00] | [1.00,1.01] | [1.00,1.01] | [0.97,0.99] |
| N healthcare facilities | 0.97 | 1.70** | 0.96 | 0.52** |
| | [0.51,1.82] | [1.09,2.66] | [0.63,1.45] | [0.30,0.90] |
| DPT-3 coverage | 0.07 | 0.00*** | 0.29 | 3.72 |
| | [0.00,8.42] | [0.00,0.00] | [0.01,13.98] | [0.01,2238.21] |
| Measles vaccination coverage | 0.94** | 1.05** | 1.01 | 0.88** |
| | [0.89,0.99] | [1.00,1.10] | [0.96,1.07] | [0.80,0.97] |
| Under-5 mortality | 0.62*** | 0.82* | 0.93 | 0.90 |
| | [0.48,0.81] | [0.66,1.03] | [0.74,1.16] | [0.61,1.32] |
| Underweight prevalence | 1.24*** | 0.99 | 1.10 | 0.85 |
| | [1.08,1.43] | [0.89,1.11] | [0.98,1.24] | [0.68,1.07] |
| Malaria prevalence, age 0-4 | 2.85 | 0.31** | 1.42 | 3.11 |
| | [0.58,14.00] | [0.12,0.84] | [0.52,3.86] | [0.43,22.44] |
| Female education, age 20-24 | 2.81*** | 2.38*** | 2.01*** | 0.65 |
| | [1.66,4.77] | [1.55,3.67] | [1.24,3.28] | [0.24,1.73] |
| Population density | 1.01 | 0.99** | 1.00 | 0.99 |
| | [1.00,1.02] | [0.99,1.00] | [1.00,1.01] | [0.98,1.00] |
| Distance to major road | 1.02 | 0.92*** | 0.93*** | 1.05 |
| | [0.94,1.10] | [0.88,0.97] | [0.89,0.98] | [0.94,1.17] |
| Road distance to regional capital | 0.91 | 1.03 | 1.13 | 1.94** |
| | [0.60,1.38] | [0.76,1.41] | [0.83,1.53] | [1.08,3.47] |
| N | 1,302 | 1,302 | 1,302 | 979 |

*Note:* Regression results for main analysis presented in Figure 2. AntenRec records whether a woman followed the recommended schedule of antenatal care visits, BirthMed records whether the child in focus was born in a medical facility, ChildVacc measures whether a child was fully vaccinated, and ChildGov measures whether a government clinic/hospital was used for the child. Reported coefficients are odds ratios, standard errors clustered at the level of the matched pair, 95% confidence intervals in brackets, $^{*}$ $p < 0.1$, $^{**}$ $p < 0.05$, $^{***}$ $p < 0.01$.

### H Conditional logistic regression

Table S7: Effect of violent victimization on healthcare-seeking behavior

| | (1) AntenRec | (2) BirthMed | (3) ChildVacc | (4) ChildGov |
|---|---|---|---|---|
| Victimized during armed violence | 1.14 | $1.26^{**}$ | $0.50^{***}$ | $0.17^{***}$ |
| | [0.89,1.45] | [1.01,1.59] | [0.40,0.63] | [0.10,0.28] |
| N | 530 | 602 | 636 | 248 |

*Note:* Replication of main analysis presented in Table S6 using conditional logistic regression models. AntenRec records whether a woman followed the recommended schedule of antenatal care visits, BirthMed records whether the child in focus was born in a medical facility, ChildVacc measures whether a child was fully vaccinated, and ChildGov measures whether a government clinic/hospital was used with the child. Reported coefficients are odds ratios, standard errors clustered at the level of the matched pair, 95% confidence intervals in brackets, $^{*}$ $p < 0.1$, $^{**}$ $p < 0.05$, $^{***}$ $p < 0.01$.

Figure S5: Effect of violent victimization on healthcare-seeking behavior, using conditional logistic regression models

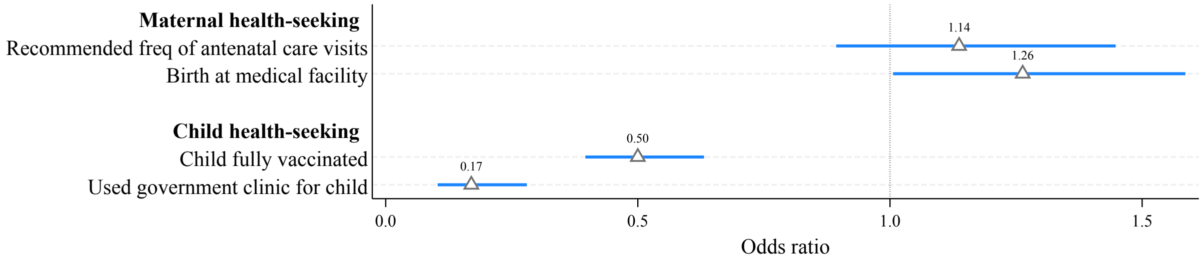


*Note:* Replication of main analysis presented in Figure 2 using conditional logistic regression models. Also see Table S7. Coefficient plots from a regression of indicators of health seeking for maternal and child health on violent victimization. Reported coefficients are odds ratios after logistic regression using the matched sample of victimized and non-victimized individuals. Markers are point estimates, lines 95% confidence intervals. Standard errors clustered at the level of the matched pair.

### I Consequences for child health

Table S8: Effect of violent victimization on child health outcomes

| | Unconditional logit | | Conditional logit | |
|---|---|---|---|---|
| | (1) BornIssue | (2) ChildIssue | (3) BornIssue | (4) ChildIssue |
| Victimized during armed violence | 5.47*** | 1.48*** | 6.00*** | 1.36** |
| | [2.64,11.34] | [1.10,1.99] | [2.96,12.15] | [1.04,1.77] |
| Female | 0.64 | 0.94 | | |
| | [0.34,1.18] | [0.64,1.37] | | |
| Age | 1.00 | 0.97*** | | |
| | [0.96,1.03] | [0.95,0.98] | | |
| Education | 0.97 | 1.14** | | |
| | [0.79,1.20] | [1.03,1.25] | | |
| Number of children | 1.02 | 1.12*** | | |
| | [0.90,1.15] | [1.06,1.18] | | |
| Child female | 0.75 | 0.90 | | |
| | [0.43,1.30] | [0.67,1.21] | | |
| Age of child | 1.30** | 1.12* | | |
| | [1.06,1.60] | [0.99,1.26] | | |
| Muslim | 8.10*** | 0.85 | | |
| | [2.73,24.08] | [0.47,1.52] | | |
| Walking time to closest health facility | 1.01*** | 1.01*** | | |
| | [1.00,1.02] | [1.00,1.02] | | |
| N healthcare facilities | 1.51 | 1.00 | | |
| | [0.63,3.60] | [0.65,1.52] | | |
| DPT-3 coverage | 0.00 | 0.00*** | | |
| | [0.00,6.01] | [0.00,0.11] | | |
| Measles vaccination coverage | 1.14*** | 1.00 | | |
| | [1.04,1.25] | [0.94,1.05] | | |
| Under-5 mortality | 1.51 | 1.11 | | |
| | [0.92,2.48] | [0.86,1.44] | | |
| Underweight prevalence | 0.71*** | 0.81*** | | |
| | [0.57,0.89] | [0.70,0.93] | | |
| Malaria prevalence, age 0-4 | 0.68 | 1.30 | | |
| | [0.14,3.44] | [0.45,3.78] | | |
| Female education, age 20-24 | 0.50 | 1.22 | | |
| | [0.17,1.41] | [0.74,2.01] | | |
| Population density | 1.00 | 0.99** | | |
| | [0.98,1.01] | [0.98,1.00] | | |
| Distance to major road | 0.96 | 0.89*** | | |
| | [0.87,1.06] | [0.84,0.95] | | |
| Road distance to regional capital | 1.45 | 1.02 | | |
| | [0.81,2.60] | [0.70,1.50] | | |
| N | 1,302 | 1,302 | 126 | 448 |

*Note:* Effect of violent victimization on child health outcomes. BornIssue records whether a child was born with a health issue (was premature; had lower-than-normal birthweight; or had other health issue) or was born healthy, ChildIssue records whether the child had one an health issue (breathing or other respiratory problems; eating or swallowing problems; stomach/intestinal problems, constipation, or diarrhea; repeated or chronic physical pain, including headaches or other back or body pain; problems using their hands or legs; problems with coordination or moving around; toothaches; deafness or problem with hearing; blindness or problem seeing; another, different health problem) during the past 12 months, or no health issue. Models 1–2 are unconditional logistic regressions, Models 3–4 are conditional logistic regressions. Reported coefficients are odds ratios, standard errors clustered at the level of the matched pair, 95% confidence intervals in brackets, $^{*}$ $p < 0.1$, $^{**}$ $p < 0.05$, $^{***}$ $p < 0.01$.

### J Robustness checks

Table S9: Effect of violent victimization on healthcare-seeking behavior, female respondents only

| | (1)<br>AntenRec | (2)<br>BirthMed | (3)<br>ChildVacc | (4)<br>ChildGov |
|---|---|---|---|---|
| Victimized during armed violence | 1.25 | 1.10 | 0.46*** | 0.20*** |
| | [0.85,1.82] | [0.81,1.49] | [0.33,0.65] | [0.11,0.39] |
| Age | 1.01 | 1.03*** | 1.01 | 0.97 |
| | [0.98,1.03] | [1.01,1.05] | [0.99,1.04] | [0.93,1.02] |
| Education | 1.01 | 1.11* | 1.18*** | 1.37*** |
| | [0.87,1.19] | [1.00,1.23] | [1.05,1.33] | [1.15,1.63] |
| Number of children | 0.92** | 0.85*** | 1.03 | 1.32** |
| | [0.85,0.99] | [0.79,0.92] | [0.96,1.10] | [1.07,1.63] |
| Child female | 1.31 | 0.91 | 0.79 | 1.07 |
| | [0.90,1.90] | [0.67,1.22] | [0.57,1.09] | [0.65,1.77] |
| Age of child | 0.88 | 0.98 | 1.97*** | 1.28* |
| | [0.75,1.03] | [0.86,1.11] | [1.65,2.34] | [0.99,1.67] |
| Muslim | 0.78 | 1.25 | 0.32*** | 0.17** |
| | [0.42,1.47] | [0.75,2.08] | [0.18,0.55] | [0.04,0.68] |
| Walking time to closest health facility | 0.99*** | 1.00 | 1.01* | 0.97*** |
| | [0.98,0.99] | [0.99,1.01] | [1.00,1.02] | [0.96,0.99] |
| N healthcare facilities | 0.95 | 1.75** | 0.86 | 0.64 |
| | [0.42,2.18] | [1.08,2.83] | [0.53,1.42] | [0.34,1.20] |
| DPT-3 coverage | 0.10 | 0.00*** | 0.30 | 20.80 |
| | [0.00,31.60] | [0.00,0.00] | [0.00,22.25] | [0.02,28367.68] |
| Measles vaccination coverage | 0.98 | 1.06** | 1.03 | 0.88** |
| | [0.92,1.04] | [1.01,1.12] | [0.97,1.09] | [0.79,0.99] |
| Under-5 mortality | 0.55*** | 0.78* | 0.94 | 0.84 |
| | [0.40,0.75] | [0.60,1.01] | [0.73,1.22] | [0.55,1.29] |
| Underweight prevalence | 1.25*** | 0.99 | 1.08 | 0.84 |
| | [1.06,1.46] | [0.86,1.12] | [0.94,1.24] | [0.64,1.10] |
| Malaria prevalence, age 0-4 | 1.11 | 0.32* | 1.19 | 2.60 |
| | [0.18,6.84] | [0.10,1.00] | [0.37,3.79] | [0.32,21.08] |
| Female education, age 20-24 | 2.13** | 2.14*** | 1.90** | 0.51 |
| | [1.15,3.94] | [1.30,3.53] | [1.08,3.36] | [0.17,1.53] |
| Population density | 1.00 | 0.99** | 1.00 | 0.99 |
| | [0.99,1.02] | [0.98,1.00] | [1.00,1.01] | [0.98,1.00] |
| Distance to major road | 1.03 | 0.91*** | 0.94** | 1.05 |
| | [0.95,1.13] | [0.86,0.96] | [0.89,0.99] | [0.95,1.17] |
| Road distance to regional capital | 0.89 | 1.04 | 1.01 | 1.95* |
| | [0.54,1.45] | [0.73,1.49] | [0.72,1.40] | [1.00,3.84] |
| N | 1,000 | 1,000 | 1,000 | 761 |

*Note:* Replication of main analysis presented in Table S6 restricting the sample to female respondents only, i.e., subsetting to female respondents. AntenRec records whether a woman followed the recommended schedule of antenatal care visits, BirthMed records whether the child in focus was born in a medical facility, ChildVacc measures whether a child was fully vaccinated, and ChildGov measures whether a government clinic/hospital was used with the child. Reported coefficients are odds ratios, standard errors clustered at the level of the matched pair, 95% confidence intervals in brackets, * $p < 0.1$, ** $p < 0.05$, *** $p < 0.01$.

Table S10: Effect of violent victimization on healthcare-seeking behavior, small-area fixed effects model

| | (1)<br>AntenRec | (2)<br>BirthMed | (3)<br>ChildVacc | (4)<br>ChildGov |
|---|---|---|---|---|
| Victimized during armed violence | 0.83 | 1.16 | 0.44*** | 0.24*** |
| | [0.55,1.25] | [0.83,1.62] | [0.31,0.62] | [0.12,0.48] |
| Female | 1.09 | 1.15 | 1.70** | 0.47 |
| | [0.65,1.82] | [0.73,1.80] | [1.06,2.70] | [0.17,1.33] |
| Age | 1.01 | 1.01 | 1.02* | 1.00 |
| | [0.99,1.04] | [0.99,1.04] | [1.00,1.04] | [0.96,1.04] |
| Education | 1.04 | 1.13** | 1.20*** | 1.31** |
| | [0.90,1.20] | [1.00,1.27] | [1.07,1.36] | [1.02,1.68] |
| Number of children | 0.91*** | 0.96 | 0.98 | 1.16*** |
| | [0.84,0.97] | [0.90,1.01] | [0.92,1.05] | [1.04,1.30] |
| Child female | 1.16 | 0.99 | 0.77 | 1.21 |
| | [0.80,1.67] | [0.74,1.33] | [0.56,1.06] | [0.65,2.26] |
| Age of child | 0.91 | 1.00 | 2.16*** | 1.28 |
| | [0.78,1.05] | [0.89,1.13] | [1.84,2.52] | [0.94,1.74] |
| Muslim | 0.82 | 1.10 | 0.69 | 0.95 |
| | [0.42,1.62] | [0.63,1.93] | [0.36,1.32] | [0.20,4.39] |
| Walking time to closest health facility | 0.99 | 1.00 | 1.02** | 1.00 |
| | [0.98,1.00] | [0.99,1.01] | [1.00,1.03] | [0.98,1.03] |
| N | 702 | 955 | 995 | 382 |
| 500×500 m FEs | Yes | Yes | Yes | Yes |

*Note:* Replication of main analysis presented in Table S6 including small-area (500×500 m) fixed effects. AntenRec records whether a woman followed the recommended schedule of antenatal care visits, BirthMed records whether the child in focus was born in a medical facility, ChildVacc measures whether a child was fully vaccinated, and ChildGov measures whether a government clinic/hospital was used with the child. Reported coefficients are odds ratios, standard errors clustered at the level of the matched pair, 95% confidence intervals in brackets, $^{*}$ $p < 0.1$, $^{**}$ $p < 0.05$, $^{***}$ $p < 0.01$.

Table S11: Effect of violent victimization on healthcare-seeking behavior, female respondents only

| | (1)<br>AntenRec | (2)<br>BirthMed | (3)<br>ChildVacc | (4)<br>ChildGov |
|---|---|---|---|---|
| Victimized during armed violence | 1.18 | 1.24** | 0.45*** | 0.24*** |
| | [0.94,1.48] | [1.01,1.53] | [0.36,0.56] | [0.17,0.33] |
| Female | 1.55*** | 1.28** | 1.56*** | 1.00 |
| | [1.22,1.96] | [1.03,1.60] | [1.24,1.95] | [0.68,1.47] |
| Age | 1.00 | 1.01 | 1.02*** | 0.99 |
| | [0.99,1.01] | [1.00,1.02] | [1.01,1.03] | [0.98,1.01] |
| Education | 1.15*** | 1.23*** | 1.26*** | 1.22*** |
| | [1.07,1.24] | [1.16,1.30] | [1.18,1.34] | [1.10,1.34] |
| Number of children | 0.97** | 0.97** | 0.99 | 1.18*** |
| | [0.94,0.99] | [0.94,1.00] | [0.96,1.01] | [1.10,1.25] |
| Muslim | 0.60*** | 1.11 | 0.47*** | 0.59 |
| | [0.41,0.88] | [0.81,1.53] | [0.34,0.66] | [0.29,1.20] |
| Child female | 1.15 | 0.99 | 0.81** | 1.16 |
| | [0.95,1.39] | [0.85,1.17] | [0.69,0.96] | [0.91,1.49] |
| Age of child | 0.94 | 1.02 | 1.61*** | 1.14** |
| | [0.87,1.02] | [0.95,1.09] | [1.49,1.74] | [1.01,1.29] |
| Walking time to closest health facility | 0.99*** | 1.00 | 1.00* | 0.99*** |
| | [0.99,0.99] | [0.99,1.00] | [0.99,1.00] | [0.98,0.99] |
| N healthcare facilities | 0.76* | 1.51*** | 0.90 | 0.69*** |
| | [0.57,1.01] | [1.19,1.90] | [0.71,1.15] | [0.52,0.91] |
| DPT-3 coverage | 0.12 | 0.00*** | 0.32 | 0.22 |
| | [0.01,2.46] | [0.00,0.01] | [0.02,4.52] | [0.00,16.76] |
| Measles vaccination coverage | 0.98 | 1.07*** | 1.05*** | 0.87*** |
| | [0.94,1.01] | [1.04,1.11] | [1.02,1.09] | [0.83,0.92] |
| Under-5 mortality | 0.72*** | 1.15** | 0.84** | 0.87 |
| | [0.61,0.86] | [1.01,1.31] | [0.73,0.96] | [0.70,1.07] |
| Underweight prevalence | 1.17*** | 0.99 | 1.02 | 1.00 |
| | [1.08,1.28] | [0.93,1.06] | [0.95,1.10] | [0.87,1.14] |
| Malaria prevalence, age 0-4 | 4.75*** | 0.25*** | 0.20*** | 3.33** |
| | [1.68,13.38] | [0.14,0.43] | [0.11,0.35] | [1.09,10.17] |
| Female education, age 20-24 | 2.20*** | 2.37*** | 1.11 | 1.60* |
| | [1.58,3.09] | [1.81,3.09] | [0.82,1.49] | [0.94,2.74] |
| Population density | 1.00 | 0.99*** | 1.00 | 0.99** |
| | [1.00,1.00] | [0.99,0.99] | [1.00,1.00] | [0.99,1.00] |
| Distance to major road | 1.00 | 0.97*** | 0.98* | 1.07 |
| | [0.95,1.04] | [0.94,0.99] | [0.95,1.00] | [0.98,1.17] |
| Road distance to regional capital | 0.81** | 0.84** | 1.16* | 2.08*** |
| | [0.66,1.00] | [0.72,0.98] | [0.98,1.36] | [1.44,3.00] |
| N | 3,006 | 3,006 | 3,006 | 2,196 |

*Note:* Replication of main analysis presented in Table S6 using the unadjusted/full sample, i.e., not using propensity score matching. AntenRec records whether a woman followed the recommended schedule of antenatal care visits, BirthMed records whether the child in focus was born in a medical facility, ChildVacc measures whether a child was fully vaccinated, and ChildGov measures whether a government clinic/hospital was used with the child. Reported coefficients are odds ratios, standard errors clustered at the level of the matched pair, 95% confidence intervals in brackets, $^{*}$ $p < 0.1$, $^{**}$ $p < 0.05$, $^{***}$ $p < 0.01$.

Table S12: Direct and witnessed victimization

| | (1) AntenRec | (2) BirthMed | (3) ChildVacc | (4) ChildGov |
|---|---|---|---|---|
| **Panel A. Victimization (all types)** | | | | |
| Victimized during armed violence | 1.14 | 1.26** | 0.50*** | 0.17*** |
| | [0.89,1.45] | [1.01,1.59] | [0.40,0.63] | [0.10,0.28] |
| N | 530 | 602 | 636 | 248 |
| **Panel B. Direct/personal victimization** | | | | |
| Personally victimized | 1.14 | 1.36 | 0.32*** | 0.05*** |
| | [0.69,1.90] | [0.83,2.21] | [0.19,0.52] | [0.01,0.20] |
| N | 164 | 176 | 220 | 94 |
| **Panel C. Indirect/witnessed violence** | | | | |
| Observed violence | 0.86 | 0.53 | 0.41** | 2.00 |
| | [0.40,1.85] | [0.24,1.13] | [0.19,0.89] | [0.18,22.06] |
| N | 164 | 176 | 220 | 94 |

*Note:* Estimated effect of different types of victimization. Panel A shows the overall estimates, as in Table S7 above; Panel B restricts the exposure to direct/personal victimization (physical assault, abduction, or sexual violence); Panel C restricts exposure to indirect/witnessed victimization (seeing someone killed or losing a household member. Analysis using conditional logistic regression with paired victimized/non-victimized cases to ensure consistent matches. Reported coefficients are odds ratios with 95% confidence intervals in brackets. $^{*}$ $p < 0.1$, $^{**}$ $p < 0.05$, $^{***}$ $p < 0.01$.

### K Direct effect on mediators

Table S13: Effect of violent victimization on mediators

| | Unconditional logit | | | Conditional logit | | |
|---|---|---|---|---|---|---|
| | (1) FearVict | (2) PTSD | (3) DistrustMed | (4) FearVict | (5) PTSD | (6) DistrustMed |
| Victimized during armed violence | 1.22* | 2.23*** | 14.54*** | 1.26** | 2.05*** | 11.46*** |
| | [0.97,1.54] | [1.73,2.88] | [10.03,21.08] | [1.00,1.57] | [1.61,2.62] | [7.79,16.87] |
| Female | 2.03*** | 0.59*** | 0.97 | | | |
| | [1.46,2.81] | [0.43,0.82] | [0.64,1.47] | | | |
| Age | 1.00 | 0.99 | 1.00 | | | |
| | [0.98,1.02] | [0.98,1.01] | [0.98,1.02] | | | |
| Education | 0.81*** | 1.15*** | 0.95 | | | |
| | [0.74,0.88] | [1.06,1.25] | [0.86,1.05] | | | |
| Number of children | 0.94** | 1.02 | 0.90*** | | | |
| | [0.89,1.00] | [0.98,1.07] | [0.84,0.95] | | | |
| Child female | 0.89 | 0.88 | 0.98 | | | |
| | [0.71,1.12] | [0.68,1.13] | [0.73,1.33] | | | |
| Age of child | 0.83*** | 1.18*** | 0.90* | | | |
| | [0.75,0.91] | [1.06,1.32] | [0.80,1.02] | | | |
| Muslim | 0.92 | 1.24 | 3.32*** | | | |
| | [0.61,1.39] | [0.82,1.89] | [1.92,5.75] | | | |
| Walking time to closest health facility | 1.01*** | 0.99** | 0.99*** | | | |
| | [1.01,1.02] | [0.99,1.00] | [0.98,1.00] | | | |
| N healthcare facilities | 0.76 | 0.94 | 1.93*** | | | |
| | [0.51,1.13] | [0.63,1.41] | [1.25,3.00] | | | |
| DPT-3 coverage | 0.25 | 0.02** | 0.46 | | | |
| | [0.01,12.31] | [0.00,0.96] | [0.00,44.42] | | | |
| Measles vaccination coverage | 1.03 | 0.99 | 1.05 | | | |
| | [0.98,1.08] | [0.95,1.04] | [0.99,1.11] | | | |
| Under-5 mortality | 1.22* | 1.08 | 0.94 | | | |
| | [0.99,1.52] | [0.87,1.34] | [0.73,1.22] | | | |
| Underweight prevalence | 0.98 | 0.86*** | 1.04 | | | |
| | [0.87,1.09] | [0.77,0.96] | [0.91,1.19] | | | |
| Malaria prevalence, age 0-4 | 16.55*** | 1.11 | 0.01*** | | | |
| | [4.64,58.96] | [0.42,2.95] | [0.00,0.07] | | | |
| Female education, age 20-24 | 1.12 | 1.22 | 0.96 | | | |
| | [0.73,1.73] | [0.79,1.86] | [0.57,1.61] | | | |
| Population density | 1.01*** | 0.99** | 0.99*** | | | |
| | [1.00,1.02] | [0.99,1.00] | [0.98,0.99] | | | |
| Distance to major road | 0.95* | 1.00 | 1.08 | | | |
| | [0.89,1.01] | [0.95,1.05] | [0.98,1.18] | | | |
| Road distance to regional capital | 0.92 | 1.41** | 1.16 | | | |
| | [0.69,1.24] | [1.05,1.90] | [0.80,1.67] | | | |
| N | 1,302 | 1,302 | 1,302 | 618 | 586 | 698 |

*Note:* Effect of violent victimization on proposed mediators. Models 1–3 are unconditional logistic regressions, Models 4–6 are conditional logistic regressions. FearVict records respondents reporting reporting fear of future violence, PTSD measures respondents suffering from post-traumatic stress disorder, and DistrustMed records whether respondents report a loss of trust in healthcare workers. Reported coefficients are odds ratios, robust standard errors with 95% confidence intervals in brackets, $^{*}$ $p < 0.1$, $^{**}$ $p < 0.05$, $^{***}$ $p < 0.01$.

### L Survey question wording

Table S14: Question wording for victimization, main outcomes, and mediators

| Variable Name | Question Text | Answer Options |
|---|---|---|
| **Violent victimization** | | |
| Affected Organized Violence | In the last 3 years, have you or any member of your household been directly affected by organized violence? | 1 Yes, I have been directly affected several times; 2 Yes, I have been directly affected once or twice; 3 No, I have never been affected by an extremist group or armed bandits; 4 Not sure; 5 Rather not answer |
| Mind Sharing Violence | Do you mind sharing what happened to you or your household member? | 1 ...was/were physically assaulted; 2 ...had a household member killed; 3 ...was/were kidnapped/abducted; 4 ...witnessed someone else being killed; 5 ...was/were threatened with violence; 6 ...experienced sexual violence; 7 ...was/were forced to leave our home; 8 ...lived in constant fear of attack; 9 ...was/were forcefully recruited by an armed group; 10 Other; 11 Don't want to say |
| Role State Security Forces | What was the role of state security forces (the police, the army) in this experience of violence? | 1 They perpetrated it; 2 They helped to push the perpetrators back; 3 They did very little/nothing about it; 4 Don't know/don't want to say |
| Help After Violence | After this experience, did you receive help from official agencies, like the (local) government, police, or the health service? | 1 Police checked on me/us or followed up; 2 Received material support (e.g., food, shelter) from local government; 3 Medical services reached out to me; 4 Received medical help after visiting a healthcare facility; 5 No, did not receive help; 6 Don't want to say |
| Fear Becoming Victim | How much do you personally fear becoming a victim of violence (again) in the near future? | 1 Very much; 2 Somewhat; 3 Not very much; 4 Not at all |
| **Maternal healthcare-seeking** | | |
| Institutionalized Birth | Where was [Name Child] birthed? | 1 In a health facility; 2 At home; 3 At traditional birth attendant's place; 4 Not sure |
| Antenatal Care | Did you/your wife get any antenatal care during your pregnancy with [Name Child]? | 1 Yes; 2 No; 3 Not sure |

*Continued on next page*

*Continued from previous page*

| Variable Name | Question Text | Answer Options |
|---|---|---|
| Visits Antenatal Care | How often did you go for antenatal visits or were visited by a nurse or doctor to receive antenatal care during the last trimester (the last 3 months) of your pregnancy? | 1 Weekly; 2 Every 2 weeks; 3 Once per Month; 5 Only once or twice; 6 Did never go/ was never visited |
| NGO support | During pregnancy and since [Name Child] was born: did you/your wife receive medical support from a local or international NGO? | 1 No, no medical help from either; 2 Yes, medical help from a local NGO; 3 Yes, medical help from an international NGO; 4 Yes, medical help from both local and international NGOs; 5 Don't know |
| Experience Seeking Healthcare | Thinking about your experiences with seeking healthcare for your children or for you/your wife over the last 3 years. Which of the following have you experienced? | 1 Arrived at clinic/hospital, but no healthcare worker was there to help us; 2 Received care for injuries inflicted by violence (e.g., bullet wounds or cuts); 3 Was at health facility that lacked even basic supplies, such as clean water; 4 Feared for my security while staying at a health facility; 5 Did not know healthcare workers well because I am new in this area; 6 Worried that healthcare facilities are unduly influenced by the government; 7 Had care or medicine provided for free by an NGO; 8 Was impressed by the courage of the healthcare workers |
| **Child healthcare-seeking** | | |
| Usual Health Facility Child | Where do you mostly go if your children are sick? | 1 Private clinic/hospital; 2 Government clinic/hospital; 3 Traditional healer; 4 NGO clinic; 5 Other; 6 None |
| Use Health Facility Child | Have you used a health facility in the last 6 months for your children? | 1 Yes; 2 No; 3 Not sure |
| Use Government Clinic Child | Which type of facility did you visit? | 1 Private clinic/hospital; 2 Government clinic; 3 Traditional healer; 4 Church/mosque clinic; 5 NGO clinic; 6 Other |
| **Child health** | | |
| Health Child Born | When [Name Child] was born, s/he. . . | 1 ...was a healthy baby; 2 ...was premature; 3 ...had a lower-than-normal birthweight; 4 ...was stillborn; 5 ...had another health issue; 6 [DON'T READ] Not sure |

*Continued on next page*

*Continued from previous page*

| **Variable Name** | **Question Text** | **Answer Options** |
|---|---|---|
| Health Issues Child | Has [Name Child] had health issues during the past 12 months? | 1 Breathing or other respiratory problems (such as wheezing or shortness of breath); 2 Eating or swallowing because of a health condition; 3 Digesting food, including stomach/intestinal problems, constipation, or diarrhea; 4 Repeated or chronic physical pain, including headaches or other back or body pain; 5 Using their hands or legs; 6 Coordination or moving around; 7 Toothaches; 8 Deafness or problem with hearing; 9 Blindness or problem seeing; 10 Others different from the above; 11 None of the above |
| **Trust** | | |
| Trust Government | How much do you trust the national government? | 1 Not at all; 2 Just a little; 3 Somewhat; 4 A lot; 5 [DON'T READ] Don't know |
| Change Trust Government | Over the past 3 years, how has your trust in the national government changed, if at all? | 1 My trust in the national government decreased; 2 My trust in the national government increased; 3 My level of trust in the national government remained about the same; 4 [DON'T READ] Not sure/Don't know |
| Trust Security Forces | How much do you trust the state security forces, such as the army and the police? | 1 Not at all; 2 Just a little; 3 Somewhat; 4 A lot; 5 Don't know/Haven't heard enough |
| Change Trust Security Forces | Over the past 3 years, how has your trust in the security forces changed, if at all? | 1 My trust in the security forces decreased; 2 My trust in the security forces increased; 3 My level of trust in the security forces remained about the same; 4 [DON'T READ] Not sure/Don't know |
| Trust Healthcare Workers | How much do you trust healthcare workers (such as doctors, nurses, etc.) and health clinics? | 1 Not at all; 2 Just a little; 3 Somewhat; 4 A lot; 5 [DON'T READ] Don't know |
| Change Trust Healthcare Workers | Over the past 3 years, how has your trust in healthcare workers and health clinics changed, if at all? | 1 My trust in healthcare workers and health clinics decreased; 2 My trust in healthcare workers and health clinics increased; 3 My level of trust in healthcare workers and health clinics remained about the same; 4 [DON'T READ] Not sure/Don't know |
| **PCL8-5 items** | | |
| Memories | Repeated, disturbing, and unwanted memories of the stressful experience? | 1 Not at all; 2 A little bit; 3 Moderately; 4 Quite a bit; 5 Extremely |

*Continued on next page*

*Continued from previous page*

| Variable Name | Question Text | Answer Options |
|---|---|---|
| Feeling Upset | Feeling very upset when something reminded you of the stressful experience? | 1 Not at all; 2 A little bit; 3 Moderately; 4 Quite a bit; 5 Extremely |
| Avoiding Memories | Avoiding memories, thoughts, or feelings related to the stressful experience? | 1 Not at all; 2 A little bit; 3 Moderately; 4 Quite a bit; 5 Extremely |
| Avoiding Reminders | Avoiding external reminders of the stressful experience? | 1 Not at all; 2 A little bit; 3 Moderately; 4 Quite a bit; 5 Extremely |
| Strong Negative Beliefs | Having strong negative beliefs about yourself, other people, or the world? | 1 Not at all; 2 A little bit; 3 Moderately; 4 Quite a bit; 5 Extremely |
| Loss Of Interest | Loss of interest in activities that you used to enjoy? | 1 Not at all; 2 A little bit; 3 Moderately; 4 Quite a bit; 5 Extremely |
| Feeling Jumpy | Feeling jumpy or easily startled? | 1 Not at all; 2 A little bit; 3 Moderately; 4 Quite a bit; 5 Extremely |
| Difficulty Concentrating | Having difficulty concentrating? | 1 Not at all; 2 A little bit; 3 Moderately; 4 Quite a bit; 5 Extremely |
| **Individual-level controls** | | |
| Gender | What is your gender identity? | 1 Male; 2 Female; 3 Other |
| Age | What year were you born? | [Number] |
| Education | What kind of education did you receive? | 1 No formal schooling; 2 Informal schooling only (including Koranic schooling); 3 Primary school completed; 4 Secondary school/high school completed; 5 Post-secondary qualifications, other than university, e.g., a diploma or degree from a polytechnic school or college; 6 Bachelor's degree; 7 Master's degree; 8 Don't know |
| Religion | What is your religion? | 1 Christian; 2 Muslim; 3 Other; 4 None |
| Number Children | How many children do you have? | [Number] |
| Sex Child | What is the sex of [Name Child]? | 1 Male; 2 Female |
| Age Child | And how old is [Name Child]? | 1 year; 2 years; 3 years; 4 years; 5 years |
| Walking Time Health Facility | How long does it take to get to the closest health clinic or health center where you can seek help if your children are ill? | [Number of minutes] |

### References SI


[1] Erik von Elm et al. “The Strengthening the Reporting of Observational Studies in Epidemiology (STROBE) Statement: Guidelines for Reporting Observational Studies”. *The Lancet* 370.9596 (Oct. 20, 2007), pp. 1453–1457.

[2] Shenyang Guo and Mark W. Fraser. *Propensity Score Analysis: Statistical Methods and Applications*. 2. ed. Advanced Quantitative Techniques in the Social Sciences Series. Los Angeles: SAGE, 2015. 421 pp.
